# Benefits of near-universal vaccination and treatment access to manage COVID-19 burden in the United States

**DOI:** 10.1101/2023.02.08.23285658

**Authors:** Fuhan Yang, Thu Nguyen-Anh Tran, Emily Howerton, Maciej F Boni, Joseph L Servadio

## Abstract

**Background:** As we enter the fourth year of the COVID-19 pandemic, SARS-CoV-2 infections still cause high morbidity and mortality in the United States. During 2020-2022, COVID-19 was one of the leading causes of death in the United States and by far the leading cause among infectious diseases. Vaccination uptake remains low despite this being an effective burden reducing intervention. The development of COVID-19 therapeutics provides hope for mitigating severe clinical outcomes. This modeling study examines combined strategies of vaccination and treatment to reduce the burden of COVID-19 epidemics over the next decade.

**Methods:** We use a validated mathematical model to evaluate the reduction of incident cases, hospitalized cases, and deaths in the United States through 2033 under various levels of vaccination and treatment coverage. We assume that future seasonal transmission patterns for COVID-19 will be similar to those of influenza virus. We account for the waning of infection-induced immunity and vaccine-induced immunity in a future with stable COVID-19 dynamics. Due to uncertainty in the duration of immunity following vaccination or infection, we consider two exponentially-distributed waning rates, with means of 365 days (one year) and 548 days (1.5 years). We also consider treatment failure, including rebound frequency, as a possible treatment outcome.

**Results:** As expected, universal vaccination is projected to eliminate transmission and mortality. Under current treatment coverage (13.7%) and vaccination coverage (49%), averages of 89,000 annual deaths (548-day waning) and 120,000 annual deaths (365-day waning) are expected by the end of this decade. Annual mortality in the United States can be reduced below 50,000 per year with >81% annual vaccination coverage, and below 10,000 annual deaths with >84% annual vaccination coverage. Universal treatment reduces hospitalizations by 88% and deaths by 93% under current vaccination coverage. A reduction in vaccination coverage requires a comparatively larger increase in treatment coverage in order for hospitalization and mortality levels to remain unchanged.

**Conclusions:** Adopting universal vaccination and universal treatment goals in the United States will likely lead to a COVID-19 mortality burden below 50,000 deaths per year, a burden comparable to that of influenza virus.

## 1 Introduction

The COVID-19 epidemic in the United States will enter its fourth year on March 1, 2023, with 507,000 Americans dying in the first year, 433,000 in the second year, and a probable 190,000 deaths in the third year (1). The epidemic response focused initially on non-pharmaceutical interventions (NPI), treatment of severe cases, vaccine rollout in early 2021, and widespread rapid testing by late 2021(2). However, none of these interventions have allowed the US COVID-19 death rate to fall below 100 deaths per day even during non-peak periods of transmission. This would be required to lower COVID deaths to an annual number near 50,000 (<20/100,000 annual mortality incidence), comparable to a severe influenza season^2^. Such a goal is reasonable for the US, a wealthy country with advanced public and private sectors in medicine and public health. To achieve a substantial mortality reduction in COVID-19 in the next several years, the most probable path lies in our past successes at controlling vaccine-preventable diseases: actively promoting universal or near-universal vaccine coverage rather than issuing recommendations for voluntary individual vaccine uptake.

The unique challenge in long-term planning of SARS-CoV-2 vaccination is that it conforms neither to our past experience eliminating childhood vaccine preventable infection such as smallpox, polio, or measles; nor can it be modeled on our current strategy to promote voluntary influenza vaccination which is meant to reduce risk of hospitalization and death in the youngest and oldest age groups. The clinical burden of SARS-CoV-2 is concentrated in the oldest age groups where population mortality rates are several times higher than those for influenza (3,4). Thus, it is likely that past approaches of universal childhood vaccination or voluntary vaccination, i.e. without urgent recommendations and proactive planning, will not work at substantially reducing the annual COVID-19 mortality burden.

Currently, there is substantial evidence that available COVID-19 vaccines are safe and effective at reducing transmission and decreasing the severity of disease (5,6). Modeling studies have shown that increasing vaccination coverage is beneficial for population health (7,8), with some evidence in favor of targeting vulnerable populations in particular (9) and of combining vaccination with NPIs (10). Similarly, current treatments have been shown to reduce the severity of disease, the likelihood of hospitalization, and the duration of infection (11-14). This is in contrast to other respiratory viruses, where treatments are not widely available (15). Therefore - with appropriate supply, access, cost, and coverage - it may be possible to substantially reduce annual SARS-CoV-2 mortality rates in the US by therapeutic interventions in the current context of voluntary vaccination. However, fewer studies have aimed to examine how to use both vaccination and therapeutics to successfully reduce mortality. It is not known what combination of increased vaccination rates and increased access to anti-COVID therapeutics would be most cost effective, and it is likewise not known what level of coverage of each is required to reduce annual death counts to levels similar to or lower than those for influenza virus.

This study aims to evaluate the effectiveness of different strategies using antiviral medications in tandem with higher levels of vaccination to adequately reduce COVID-19 morbidity and mortality in future epidemic years. We adapted a previously validated mathematical model of SARS-CoV-2 epidemiology and clinical progression in the US to predict future burden and then implemented various strategies of vaccine and antiviral deployments. We compared effectiveness of strategies via reductions in case totals, hospitalizations, and deaths. The results of this study provide insight into the benefits of using both annual vaccines and antiviral medications to reduce long-term COVID-19 burden.

## 2 Methods

This study uses a previously published dynamical model (Fig. S1) (16-18) designed to examine COVID-19 burden in Rhode Island (RI), Massachusetts, Connecticut, and Pennsylvania. The model incorporates key aspects of SARS-CoV-2 dynamics, including asymptomatic transmission, vaccination, and age-specific (10-year age bands) risks of infection and severe outcomes. Eleven daily data streams were used for the Bayesian model fit, which showed consistency in its inference of clinical and epidemiological transition parameters across four different states and at different phases of data collection (16,17). We adapted the previously published model, calibrated the model based on its fit for RI (17), and then produced simulations for the present study.

### 2.1. Data sources

Daily data for cumulative cases, hospitalizations, and deaths are publicly available from the RI Department of Health (DOH) (19). From these, we calculated daily incident cases, hospital admissions, and deaths. To apply results from a model calibrated to RI to the entire US, we collected weekly incident cases and deaths for the US, which are publicly available from the US Centers for Disease Control and Prevention (CDC) (1), and weekly hospitalization data, which are publicly available from the Department of Health and Human Services (20). These data are available by state and were summed to represent national burden. For both RI and the US, we collected data between Mar 1, 2020 and Nov 30, 2022.

COVID-19 vaccination data for RI were assembled from RI DOH and CDC. Weekly counts of people who completed the doses of a primary series (two doses of Pfizer or Moderna vaccines or single dose of Johnson and Johnson vaccine) from RI are publicly available from RI DOH (19) from Dec 13, 2020 to Jul 30, 2022. The data afterwards are available from CDC (21) until Nov 30, 2022. Both data sources contain age-specific totals of individuals receiving a COVID-19 vaccine (either an initial series or booster). The age groups from CDC were: 0-4, 5-12, 13-17, 18-64, 65+, and these were adjusted to our 10-year age bands assuming a uniform distribution of coverage within each age band.

Monthly influenza vaccination coverage from season 2010-2011 to season 2020-2021 was collected from the CDC (22), provided by the National Immunization Survey-Flu (NIS-Flu) and the Behavioral Risk Factor Surveillance System (BRFSS). Coverage was estimated from the proportion of the US population that received an influenza vaccine based on telephone surveys conducted from October to May in each season. Coverage among children (6 months - 17 years old) is reported by NIS-Flu, and coverage among adults is reported by BRFSS.

The national and jurisdictional cumulative counts of delivered and administered therapeutics including nirmatrelvir/ritonavir (Paxlovid), molnupiravir (Lagevrio), and Tixagevimab/cilgavimab (Evusheld) are available from HHS (23). We defined treatment coverage in the current season as the coverage of Paxlovid because it is the most commonly used COVID-19 therapeutic in the US. Current treatment coverage is 13.7%, calculated from total administered doses of Paxlovid (6,279,116) and total COVID-19 cases (45,666,906) between Jan 1, 2022 and Nov 30, 2022 in the US.

### 2.2. Model adaptation

We introduced one vaccination class to the previously developed model; after receiving a vaccine, individuals are moved to this class (“Vac” in Fig. S1). Candidates for vaccination include infected individuals who are presymptomatic or asymptomatic, individuals who recently recovered or were discharged from hospital, and susceptible individuals. Recovered, discharged, and susceptible populations move into the vaccinated class, while presymptomatic and asymptomatic infected individuals continue on their normal course of infection. From December 13, 2020 - November 30, 2022, the vaccination rate was determined by the weekly number of people who completed a primary series (two doses from Pfizer or Moderna, or one dose from Johnson & Johnson) reported by RI DOH or CDC.

To account for treatment of symptomatic individuals, additional transitions were added between classes. During the 6-day infection period, symptomatic individuals who receive treatment will do so during the first three days of infection. As a rebounding effect (the relapse of symptoms or viral load within a short time after treatment) of the therapeutics has been reported (24-29), we consider three treatment outcomes: (*i*) successful treatment, where patients recover completely after treatment and move to the recovered class after day 3 of infection; (*ii*) treatment failure, which includes rebound, where patients are still symptomatic and infectious after failed treatment; and (*iii*) hospitalization, where patients are hospitalized after failed treatment. Treatment efficacy is modeled by two parameters: probability of treatment failure, and hospitalization fraction given treatment failure. We set the probability of treatment failure to 0.035 based on population studies on the rebounding effect of Paxlovid and Molnupiravir (29,30). The probability of hospitalization after failed treatment is assumed to be 88% lower when compared to no treatment (11,31). Other values for these parameters are discussed in section 3.5.

We introduced waning of infection-induced and vaccine-induced immunity starting at the beginning of the Delta period (June 2021). A number of studies have been published comparing times to reinfection among individuals who have or have not been previously vaccinated or infected (32-39) many of which align with an estimated 40% to 70% becoming susceptible to reinfection after one year (Table S4). Comparison among studies and inference on absolute measures of protection are challenging as the studies (*i*) used different controls groups and (*ii*) used cohorts that were exposed to infection during different periods of the pandemic. Nevertheless, the rates of reinfection in these studies appear to be consistent with average rates of immune waning between 365 days (one year) and 548 days (1.5 years). An exponentially-distributed 548-day waning period implies that 49% of individuals will lose immunity one year after infection or vaccination, and 74% will lose immunity after two years. A 365-day waning period implies that 63% of individuals will lose immunity one year after infection or vaccination, and 86% will lose immunity after two years. A 730-day waning period is considered in the supplementary materials, though this appears to be less consistent with available data. Model results for 365-day and 548-day waning are presented in the main text.

Parameters pertaining to clinical progression, such as probability of symptomatic infection, hospitalization, and death, were previously fit for RI using data through June 6, 2021(17); we used these values in the model for this time period. These, as well as parameters for vaccine and treatment efficacies(40), were updated based on published literature for the Delta period (June 7, 2021 - December 20, 2021) and Omicron period (December 21, 2021 onward) (41-45). Details are presented in Table S1. We assumed 95% efficacy of the annual vaccine for all periods (36).

Between June 7, 2021, and November 30, 2022, we calibrated the time-varying transmission rate to fit the daily hospitalized cases in RI such that at least 85% of the days within this period produced modeled hospitalization values within 10% of the observed hospitalization incidence. We then applied the average transmission rate parameter between May 1 and July 1, 2022 to December 1, 2022 through February 28, 2023, multiplied by a factor such that the observed deaths between March 1, 2022 and February 28, 2023 would be approximately 170,000, reflecting the probably number of deaths during this period (162,136 between March 2, 2022 and January 31, 2023) (1). To apply results from a RI-calibrated model to the entire US, we scaled model outputs using the ratio of the total cases between the US and RI during each of the four variant periods: wildtype (March 1, 2020 - March 30, 2021), Alpha (March 31, 2021 - June 6, 2021), Delta (June 7, 2021 - December 20, 2021), Omicron (December 21, 2021 onward). The US population is 330 times larger than the RI population, and US case numbers were between 220 and 240 higher than RI case numbers; this is likely due to higher population density and higher-than-average reporting in Rhode Island. The scaled-up results match the trends observed in the US (Fig. S2).

Using current coverage of vaccination and treatment, we established a “status quo” scenario, representing COVID-19 projections if vaccination and treatment use remain unchanged. We set the status quo treatment coverage at 13.7%, and the initial vaccination coverage to 49% based on the allocation of Paxlovid and the administered number of primary series and boosters during the third year of the pandemic (2022-2023) (detailed assumption of vaccine coverage are in S1 Text and Fig. S3). Current statewide levels of treatment and vaccination are available in Fig. S4 and Fig. S5.

### 2.3. Projecting future disease burden

We adjusted the transmission levels in the model for future projections based on influenza dynamics in the US. Seasonal wintertime forcing in transmissibility was introduced starting in season 2023-2024, where transmission increases by up to 20 percent in a sinusoidal curve between October and February, estimated from influenza transmission during winter in the northeastern US (46). The baseline transmission rate after March 1, 2023 was assumed to be the mean of the transmission parameter throughout the entire third year. We assumed monthly COVID-19 vaccination patterns would resemble those of influenza vaccinations. Age-specific proportions of influenza vaccines administered during each month were linearly interpolated to generate age-specific proportions of administered COVID-19 vaccines administered each week in a year (Details in S1.Text). The annual age-specific trend of vaccination is available in Fig. S6.

From the status quo, we generated model projections from March 1, 2023, to February 28, 2033, and recorded cumulative reported cases, hospital admissions, and deaths. We then applied changes in vaccine coverage and treatment coverage to the model to estimate burden under different strategies of vaccination and/or treatment. Because dynamics were unstable over the first two seasons (2023, 2024), averaged annual cases, hospital admissions, and deaths over 2025-2033 were used as the primary measure for comparison across strategies. We repeated analyses for two additional transmissibility scenarios, optimistic and pessimistic, referring to transmissibility equal to half and double, respectively, of that observed during the 2022-2023 year.

## 3 Results

### 3.1. Status quo projections

If current vaccination and treatment coverage (49% and 13.7%, respectively) do not change going forward, our model projects an average annual COVID-19 burden of 22.2 million cases, 742,000 hospitalizations, and 89,000 deaths, for 1.5-year immune waning, and an annual burden of 31.8 million cases, 1,045,000 hospitalizations, and 120,000 deaths for one-year immune waning. In an ‘optimistic’ scenario where future transmission rates are half of the estimated transmission rates for the 2022-2023 season, yearly burden ranges from 10.5 million to 12.5 million cases, 307,000 to 352,000 hospitalizations, and 32,000 to 35,000 deaths (across the two immune waning rates). In a ‘pessimistic’ scenario where transmission is doubled, yearly burden is estimated to be between 34.5 million and 50.6 million cases, 1.3 million and 1.9 million hospitalizations, and 176,000 and 250,000 deaths. The neutral and pessimistic scenarios both predict mortality from COVID-19 that substantially exceeds that from influenza seasons (range of annual mortality: 12,000 - 52,000 (47)) if no increases are made to vaccination or treatment coverage.

### 3.2. Increasing vaccination coverage

Assuming treatment coverage remains at current levels, increasing vaccination coverage alone reduces cases, hospitalizations, and deaths. Under neutral assumptions about future transmissibility, our model shows that annual deaths from COVID-19 during the next 5-10 years can be reduced to fewer than 50,000 (comparable to a severe influenza season) if more than 61% (548-day immune waning) or 81% (365-day immune waning) of the population is vaccinated every year. Deaths can be reduced to fewer than 10,000 if more than 68% (548-day immune waning) or 84% (365-day immune waning) of the population is vaccinated every year (Fig. 1). In these neutral scenarios, disease burden would be eliminated by the start of the 2025-2026 season if annual vaccination reaches 81% or 96% coverage, respectively. This range of thresholds underlines the important effect of immune waning when forecasting future epidemic waves of SARS-CoV-2.

**Figure 1.**
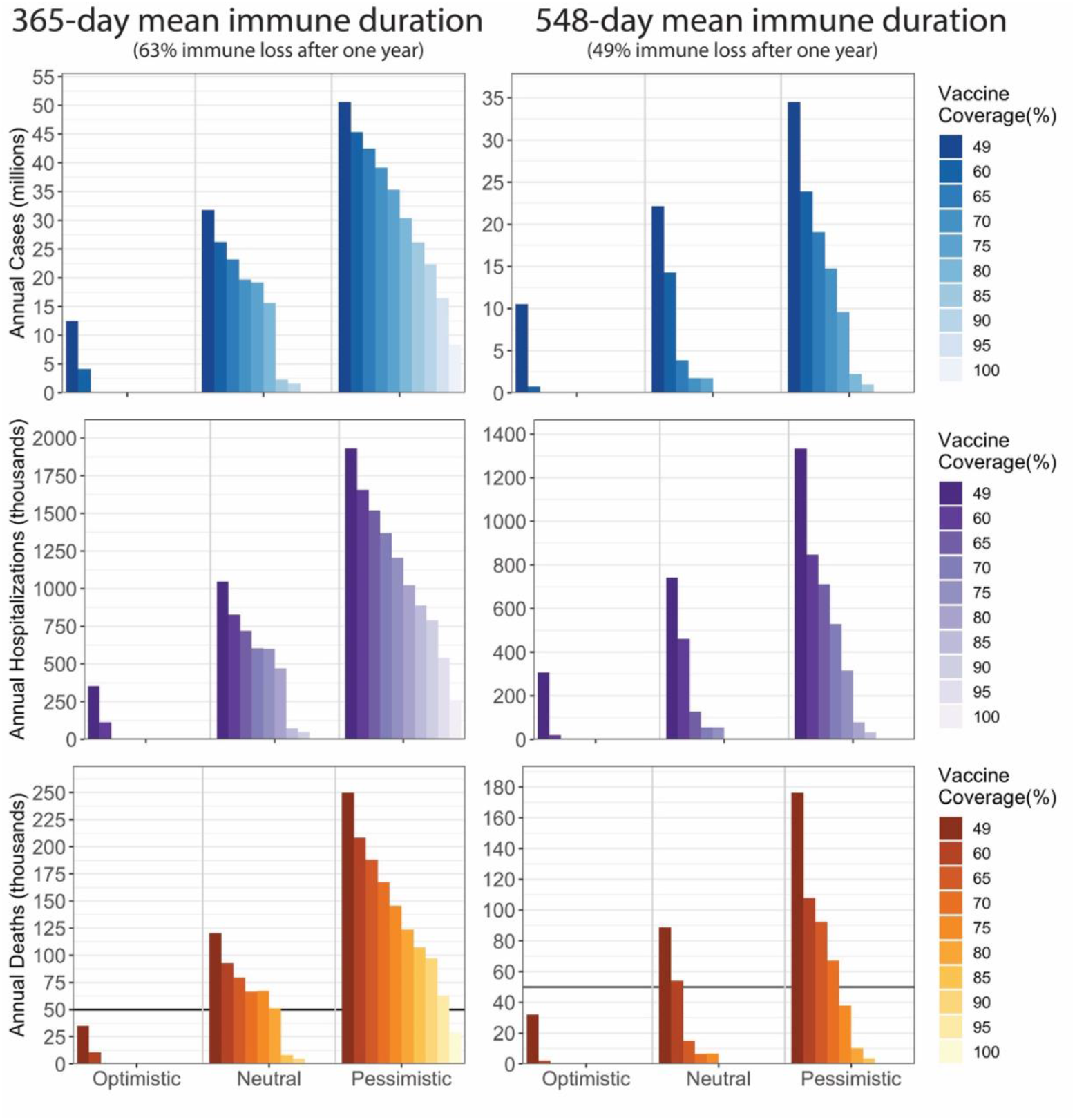
Annual cases, hospitalizations, and deaths between 2025 and 2033 under varying vaccination coverages in three transmission scenarios and two immune durations (365 days and 548 days). The current vaccination coverage is 49%. The treatment coverage through the entire period is 13.7%.

If future transmissibility is halved, current vaccination coverage (49%) would reduce deaths below 50,000 for both durations of immune waning. Increasing the coverage to 54%-61% (for the two waning rates) would reduce annual deaths below 10,000 by the 2025-2026 season, and increasing coverage to 68%-77% would eliminate mortality at the beginning of the 2025-2026 season. In the pessimistic scenario where transmissibility is doubled, even universal vaccination coverage is not projected to reduce annual mortality below 10,000 if the true mean duration of immune waning is 365 days. Annual vaccination coverage needs to reach 73%-97% to achieve fewer than 50,000 deaths per year (for the two waning rates). For 548-day immune waning, 81% coverage is needed to reduce annual deaths below 10,000 and 96% annual vaccine coverage is needed to eliminate mortality as we enter the 2025-2026 season.

### 3.3. Increasing treatment coverage

Assuming current vaccination coverage and virus transmissibility remain unchanged, treatment coverage or treatment access rates of 53% (548-day immune waning) or 66% (365-day waning) would be sufficient to reduce annual death rates below 50,000. Treatment coverages of 97% and 100%, respectively, would be required to reduce annual death rates below 10,000. Benefits of universal treatment are consistent across the three transmission scenarios and two immune durations with average reductions of 18% for cases, 88% for hospitalizations, and 93% for deaths. Increasing population-level treatment coverage reduces disease burden in a linear manner (Fig. 2) and is more effective at reducing hospitalizations and deaths than infections. We project that a 10% increase (absolute percentage points) in treatment coverage leads to an average annual reduction of 237,000 cases, 74,000 hospitalizations, and 9,400 deaths (548-day immune waning), or a reduction of 420,000 cases, 111,000 hospitalizations and 14,400 deaths (365-day immune waning). As expected, access to treatment becomes more important as vaccination coverage declines (Fig. S7) and if future transmission rates increase (Fig. 2); though the relative decrease is similar across scenarios, the absolute difference changes substantially.

**Figure 2.**
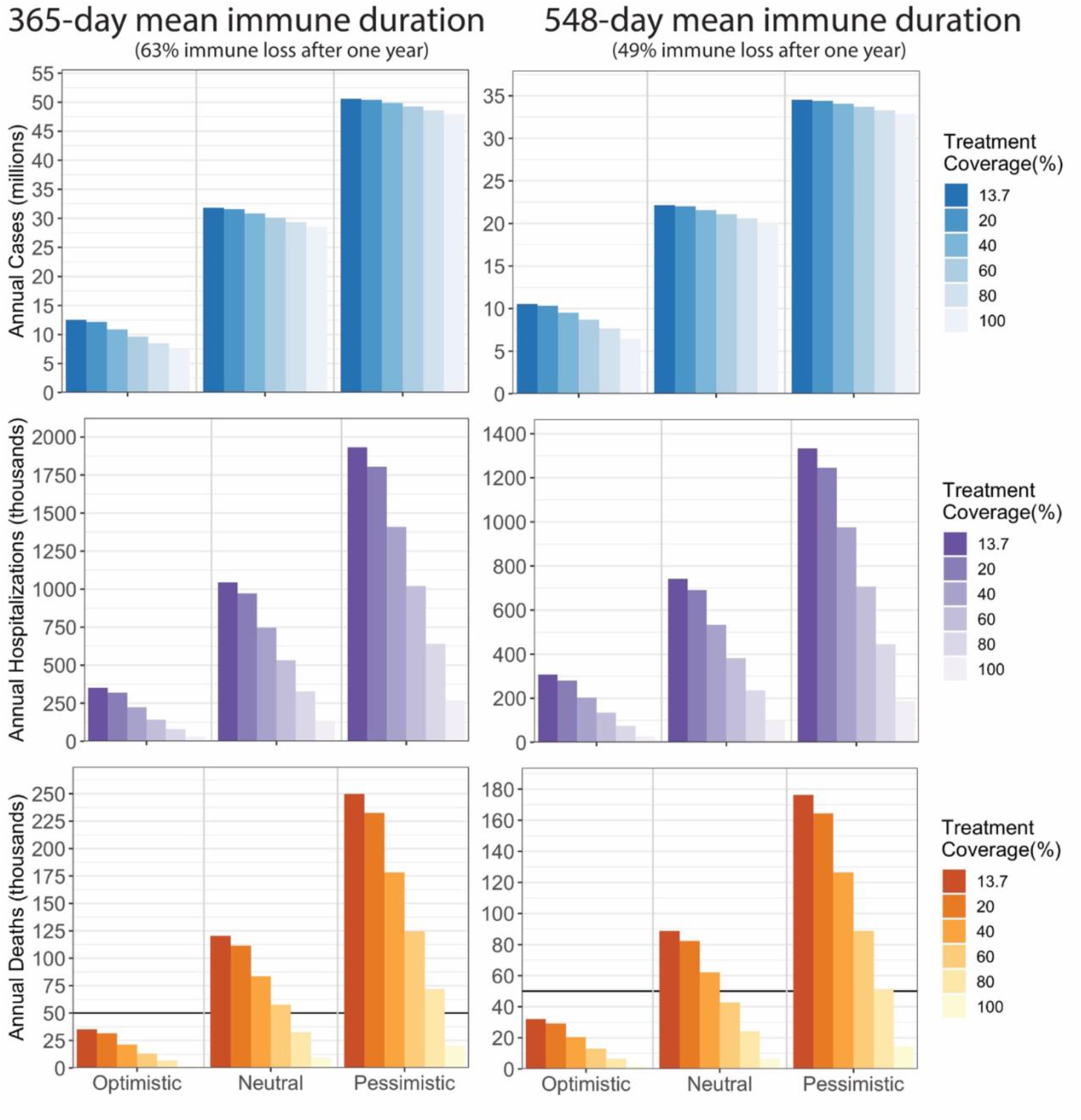
Annual cases, hospitalizations, and deaths between 2025 and 2033 under varying vaccination coverage in three transmission scenarios and two immune durations (365 days and 548 days). The current treatment coverage is 13.7%. The vaccination coverage through the entire period is 49%.

### 3.4. Combining strategies of vaccination and treatment coverage

We varied treatment and vaccine coverage simultaneously to identify combinations that drive annual death counts to more acceptable levels. For example, if vaccination coverage can only be increased to 55%, then 55% vaccine coverage and 20% treatment coverage (548-day waning) or 55% vaccine coverage and 50% treatment coverage (365-day waning) would reduce annual death counts below 50,000 (Fig. 3, Fig. S8). The accrual of public health benefits is linear with increasing treatment coverage but non-linear (accelerating) with increasing vaccination coverage. Sufficiently high vaccination rates will eliminate deaths altogether regardless of treatment coverage, but the same is not true for sufficiently high treatment access. Multiple combinations of vaccine/treatment access are projected to lead to annual death counts below 50,000; as examples, 56%/15% access, 50%/35% access, and 42%/55% access are all associated with ∼50,000 annual deaths (Fig. 3). A small drop in vaccination coverage requires a comparatively larger compensatory increase in treatment coverage to achieve the same mortality result. Similar effects of vaccination and treatment are observed in reducing hospitalization (Fig. S9).

**Figure 3.**
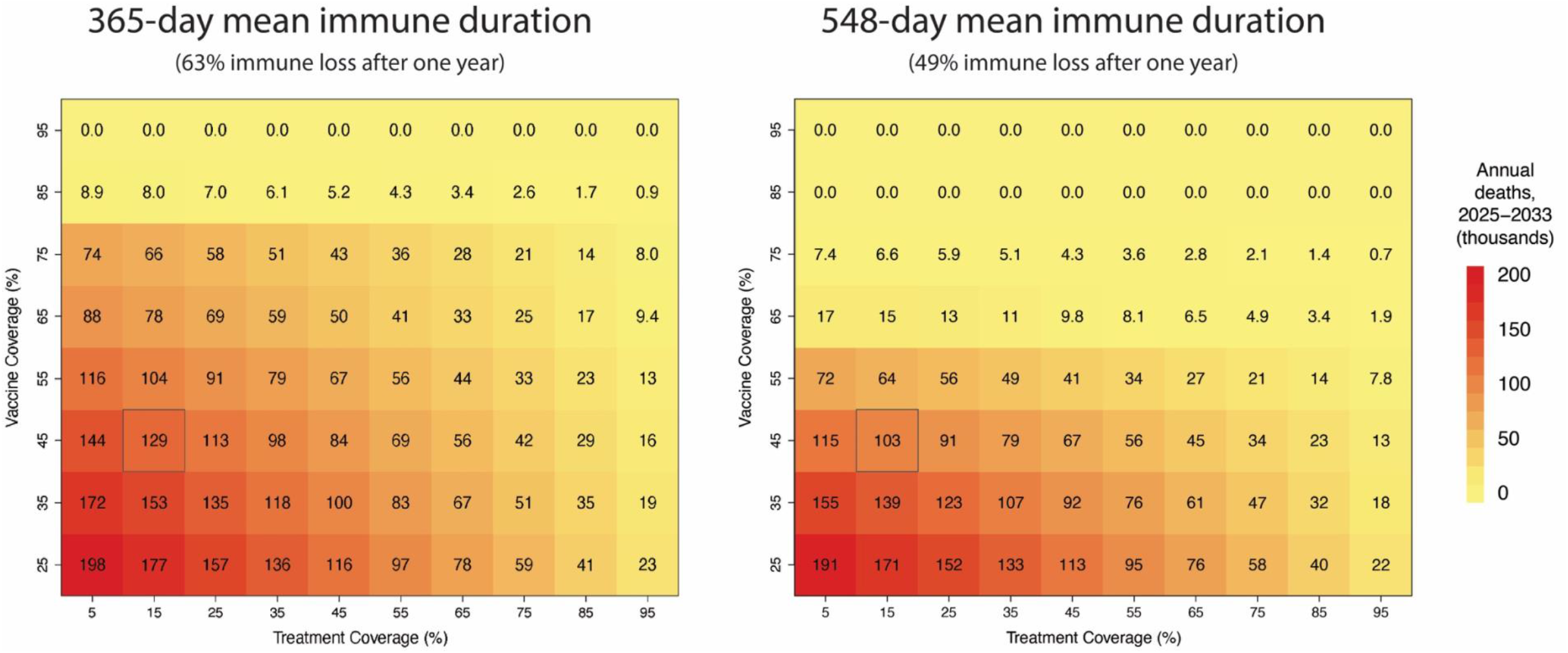
Mean annual deaths (in thousands) between 2025 and 2033 under various combinations of vaccination and treatment coverage. Two rates of immune waning are considered, one-year (left) and 1.5 years (right). The outlined cell represents the treatment and vaccination coverages closest to observed levels (49% vaccine coverage, 13.7% treatment coverage).

Current vaccine and treatment coverages by state serve as examples of achievable coverage levels at the national level (Fig. S4, Fig. S5). Additionally, this information identifies states requiring the most effort and highest resource allocation to improve coverage rates, as well as those currently pursuing strategies that would yield adequately low annual mortality. Under 548-day immune waning, only four states have current vaccination and treatment coverages that would result in annual deaths below 10,000 (if those coverages were reached nationally), and six have coverages that would achieve between 10,000 and 50,000 deaths (Fig. 4). Under 365-day immune waning, no state’s coverage would achieve under 10,000 deaths, and one state’s coverage would achieve under 50,000 deaths if applied nationally. Most states are under-vaccinated, with current coverage levels that would be associated >50,000 annual deaths nationally under 548-day waning. Even higher coverage of vaccination and treatment would be needed under 365-day immune waning (Fig. S11).

**Figure 4.**
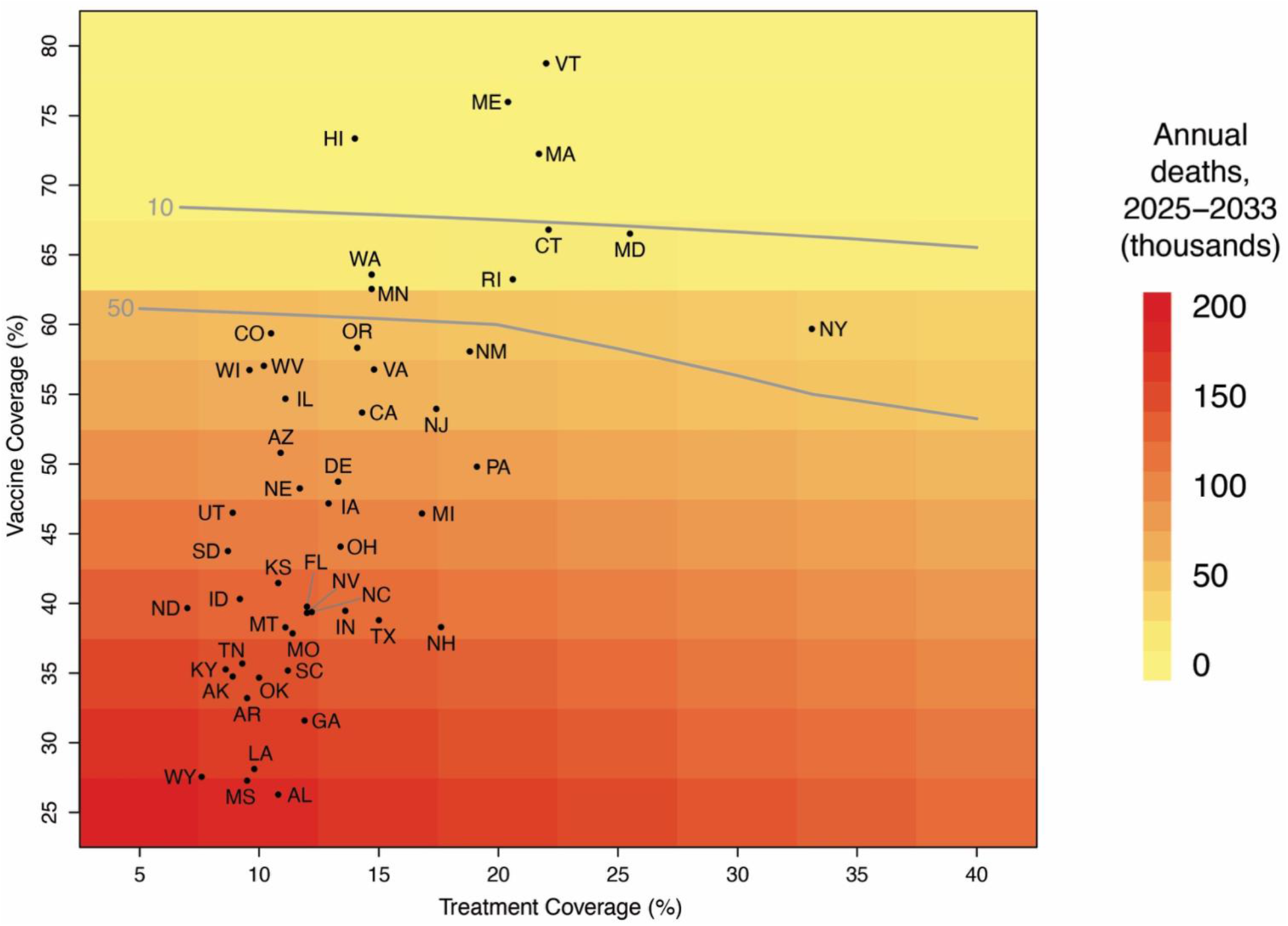
Estimated annual burden between 2025 and 2033 under 1.5-year immunity waning if implementing the current vaccination and treatment coverages of 50 states nationally (black dots). Contour lines show 10,000 and 50,000 annual deaths, representing the highest and lowest deaths from recently observed influenza seasons.

### 3.5. Potential risks of vaccine or treatment failure

To account for uncertainty in future vaccine and treatment efficacies, we examined additional scenarios to determine how model projections would change if vaccine or treatment efficacies dropped. In all three transmission scenarios, disease burden can still be eliminated after the 2025-2026 season under high coverage of a vaccine with 88% efficacy (48) (this was chosen as the efficacy of primary series of vaccinations against the Delta variant); at least 67%-76% coverage would be required under the optimistic scenario (for the two waning rates), 76%-95% under the neutral scenario, and 95% coverage under the pessimistic scenario for 548-day immune duration. For a vaccine with 50% efficacy - to provide an example of an immune-escape scenario like the one seen when the Omicron variant emerged in autumn 2021 - universal vaccination would not eliminate mortality burden (Fig. S12).

Post-treatment viral rebound is a concern for treatment uptake and efficacy, but it is currently rare, seen in <10% of patients (24-28,49). Our projections indicate that if the probability of treatment failure increases to 20%, universal treatment would still be able to reduce mortality by 85% and hospitalizations by 87%, on average, across all three transmission scenarios and both immune durations (Fig. S13). Further, if the true risk reduction for hospitalization of treatment is 50%, lower than the originally reported 88%, as indicated in recent CDC reports (12), universal treatment is projected to reduce mortality by 78% and hospitalization by 56% on average across all three transmission scenarios and two immune durations. Similarly, using a risk reduction for hospitalization of 30% (as seen in molnupiravir trials (50)) universal treatment is projected to reduce mortality by 70% and hospitalization by 41% on average across all three scenarios and two immune durations (Fig. S14).

## 4 Discussion

We evaluated the benefits of universal and near-universal vaccination and treatment at reducing COVID-19 burden in the United States using projections from a previously fit and validated mathematical model. This study contributes to current literature examining the benefits of vaccination coverage to manage COVID-19 (9,51,52) while also incorporating treatment coverage (53). This contrasts with the majority of other treatment-focused studies by examining population benefits rather than individual benefits (54,55). As expected (53,56), simulations of SARS-CoV-2 dynamics through 2033 show that high vaccination coverage and high treatment coverage are both effective means of lowering COVID-19 deaths below 50,000 deaths annually - a crucial psychological marker that will allow our public health system to state that control of SARS-CoV-2 is approximately as successful as control of influenza virus. Current (as of Nov 30, 2022) vaccine and treatment coverage levels are projected to lead to 89,000 - 120,000 deaths annually (range: 32,000 - 250,000, across various scenarios of transmission severity and immune durations), a mortality level that exceeds the most severe influenza seasons.

As we enter the fourth year of the pandemic, COVID-19-associated mortality is still the largest among all infectious pathogens in the US. The next highest annual infectious disease mortality burdens in the US are attributed to influenza virus and pneumonia (between 12,000 and 52,000 deaths annually), respiratory syncytial virus (between 5,000 and 15,000 annual deaths) (57), HIV/AIDS (∼6,000), and viral hepatitis (∼5,000) (58). Clearly, any controllable infectious pathogen that causes >100,000 annual deaths needs constant prioritization and the full attention of the Centers for Disease Control and Prevention, State Departments of Health, and federal budgeting decisions until annual death numbers can be brought down to levels comparable to other infectious diseases.

Current vaccination coverage in the US remains low for a variety of reasons including lack of access, coordinated disinformation campaigns (59,60), and vaccine hesitancy (61). Antiviral therapeutics and monoclonal antibody treatments (Table S2) offer another effective way of reducing COVID-19 hospitalizations and deaths when vaccine hesitancy may be too strong to overcome in the near term. However, as a long-term approach, it is important to remember that treatment offers direct benefits only (to the patient receiving the treatment) while vaccination provides both direct and indirect benefits by lowering the probability of (*i*) one’s own infection and (*ii*) onward infections they would have caused. The increased benefit of vaccination can be seen in Fig. 3, where increasing vaccination alone leads to greater reductions in mortality compared to similar increases in treatment, a finding echoed in other studies (53). As vaccine coverage increases, the indirect benefits of vaccination do not suffer from diminishing returns; rather, they benefit from accelerating returns (62,63). This means that every additional vaccinee lowers total COVID-19 risk more than the previous vaccinee. It is imperative that we remind the nation’s public health leadership of this basic fact of epidemiology as experience from the successful measles, smallpox, and polio vaccination campaigns has faded from memory over the past 75 years. Additionally, although NPIs have proven to be effective (64,65), they are best used as emergency measures while other interventions are insufficient or not yet available (e.g. during 2020 before COVID vaccines were approved). In a future with endemic COVID-19, vaccination and treatment are the most readily adoptable and sustainable interventions, as they do not require large-scale or long-term behavioral changes such as persistent masking, event cancellations, movement restrictions, and reduced social contacts.

The results of this study can help inform resource prioritization for vaccination and/or treatment. Prioritization can be evaluated at both regional and state levels, as vaccination and treatment coverage vary substantially across and within states (Fig. 4). Identifying states that should be prioritized for increased vaccination and/or treatment access would be an ideal way to initiate a national public health campaign intended to cover the majority of Americans with access to vaccines and COVID-19 treatments. The vertical and horizontal shifts in Figs. S9-S11 can be used to anticipate changes in total case numbers and absenteeism, strains on healthcare systems, and projected death counts if vaccination and treatment access are inadequate.

Evaluating vaccination and treatment as national coverage percentages does not account for nonuniform access to these interventions and the nonuniform risk of infection and severe outcomes that is already well known for SARS-CoV-2 transmission in the US. Prior to and throughout the COVID-19 pandemic, access to healthcare has been shown to vary substantially among various geographic and demographic groups. It is important to consider implementation strategies that are both effective and equitable (66,67). Therefore, we should be careful not to aim absolute or national-level coverage recommendations at an uncertain herd immunity threshold - a mistake made in early 2021 during the initial vaccine rollouts (17). Recommendation should instead aim for universal access and coverage for both vaccination and treatment through strong and targeted recommendations adapted to every community’s priorities and needs. Passive recommendations for voluntary vaccination have so far proven unable to move annual COVID-19 vaccination past the 50% coverage mark (68).

### 4.1. Limitations

One notable limitation in our analysis is that we assume that SARS-CoV-2 dynamics in the US will progress in accordance to mixing patterns, reporting rates, and health care access as seen in our RI-based parameterization. Inference on clinical parameters - progression rates to hospitalization, ICU care, death - and durations of infection and hospital care proved to be robust across four states (16,17) and >40 statistical fits performed during 2020 and 2021 at different stages of data completeness. This suggests that the basic clinical timeline of a COVID-19 case is similar across states and different public health systems. However, population density, mixing rates, and compliance with NPIs do vary among states and regions in the US. We attempted to account for this by scaling national case numbers to RI-case numbers, but the unaccounted heterogeneity among states and counties will certainly have an effect on the implied herd immunity levels shown in Fig. 3.

We did not incorporate population demographics into our model, preventing us from accounting for the expected pace of demographic shifts over the next ten years. The age structure of the model population remained unchanged during the entire projection period, and the model did not include natural births or deaths. Natural births introduce a population completely immune-naïve to SARS-CoV-2, while our model assumes a population that has almost certainly been previously infected or vaccinated by March 2023. This may likely lead to our burden estimates being an underestimate due to young children being more susceptible than our model indicated.

We assume that the duration of immune protection is the same after infection or vaccination; however, it is possible that the waning rates of immunity gained from vaccination and infection differ, though current studies do not appear to have conclusive answers to this question (37). Our results show that average annual mortality differs based on the two mean immune waning rates presented here, and further changes to our methodology would be needed if infection- and vaccine-induced immunity are proven to wane differently.

### 4.2. Future Directions

Increasingly, rebound reports during Paxlovid treatment are being published (24-28,49,69); however, some studies also show rebound occurring during natural infection (69-71). As rebound may be independent of treatment, our model assumes that failed treatments or rebounds have lower risks of hospitalization than that for untreated individuals. Even accounting for rebound effects, model results still indicate that high levels of treatment access will lead to substantial reductions in hospitalization and death. We considered other reductions in the risk of hospitalization after treatment (Fig. S14), as the relative reduction in hospital admissions will vary by treatment and by the control group against which comparison is done. As more data are collected on treatment efficacy, for currently circulating variants and current levels of hospitalization, it will be imperative to update current projections based on the most commonly available and prescribed drugs. As more patients receive antiviral treatments, another important concern will be the potential for emergence of drug-resistant genotypes. Mutations in SARS-CoV-2 following treatment with nirmatrelvir, a component of Paxlovid, have already been observed in the laboratory (72,73). Thus, it is important to continue monitoring outcomes of treatment efficacy studies, and policy should be updated based on any major changes in treatment profile, efficacy, or drug resistance.

The costs of various public health measures to increase vaccination or treatment access are likely to differ. The primary focus of this study was reduction of COVID-19 disease burden and not cost-effectiveness, and the same reduction in mortality achieved through modifying either treatment or vaccination is viewed as equivalent despite having potentially different financial costs. Detailed costing studies will be needed to evaluate the cost-effectiveness of various combinations of vaccine and treatment coverage in reducing morbidity and mortality.

## 5 Conclusions

A total of 190,000 Americans will have died by the end of the third year of the COVID-19 pandemic, a fact that must be viewed as a public health failure given (*i*) the availability of vaccines in 2022, (*ii*) an update to include the Omicron variant in a bivalent vaccine formulation, and (*iii*) the vaccines’ approval in June 2022 for children under five - the last age group to be vaccinated. Although it is still not clear whether the planning of new and active public health campaigns should aim to get vaccination coverage levels to 80% or 90% or higher, it is clear that the vast majority of US states are under-vaccinated and probable that >100,000 Americans will die annually from COVID if no major improvements are made in vaccine adoption. The simplest approach to narrowing this gap appears to be a recommendation for universal vaccination and a measurement each year of how effectively this recommendation (and its associated efforts and policies) is working in different states and age groups. Although it is unlikely that changes in vaccination coverage can be achieved quickly, we would strongly urge our national-level public health leadership to begin making plans for outreach and communication around universal COVID-19 vaccine coverage.

## Data Availability

All data produced and relevant code used in the present study are available online at https://github.com/Fuhan-Yang/covid-treatment-psu-cidd

## List of Abbreviations

COVID or COVID-19: Coronavirus infectious disease 2019
US: the United States
NPI: non-pharmaceutical interventions
RI: Rhode Island
DOH: Department of Health
CDC: Center for Disease Control and Prevention
HHS: Health and Human Services
BRFSS: Behavioral risk factor surveillance system
NIS-Flu: National immunization survey-flu

## Availability of data and materials

Relevant data and code for this study are available at https://github.com/Fuhan-Yang/covid-treatment-psu-cidd

## Author contributions

Conceptualization: FY, MFB, JLS

Data curation: FY, TNAT, EH, MFB

Formal analysis: FY, JLS

Methodology: FY, TNAT, EH, MFB, JLS

Software: FY, TNAT

Supervision: MFB, JLS Validation: FY

Visualization: FY, JLS

Writing - original: FY, EH, JLS

Writing - review: FY, TNAT, EH, MFB, JLS

## References

1. CDC. COVID Data Tracker [Internet]. Centers for Disease Control and Prevention. 2020 [cited 2023 Jan 23]. Available from: https://covid.cdc.gov/covid-data-tracker

2. Peeling RW, Heymann DL, Teo YY, Garcia PJ. Diagnostics for COVID-19: moving from pandemic response to control. Lancet. 2022 Feb 19;399(10326):757–68.

3. Torres C, García J, Meslé F, Barbieri M, Bonnet F, Camarda CG, et al. Identifying age-and sex-specific COVID-19 mortality trends over time in six countries. Int J Infect Dis. 2022 Dec 10;128:32–40.

4. Quandelacy TM, Viboud C, Charu V, Lipsitch M, Goldstein E. Age-and Sex-related Risk Factors for Influenza-associated Mortality in the United States Between 1997-2007. American Journal of Epidemiology. 2014 Jan 15;179(2):156–67.

5. Zheng C, Shao W, Chen X, Zhang B, Wang G, Zhang W. Real-world effectiveness of COVID-19 vaccines: a literature review and meta-analysis. Int J Infect Dis. 2022 Jan;114:252–60.

6. Lopez Bernal J, Andrews N, Gower C, Robertson C, Stowe J, Tessier E, et al. Effectiveness of the Pfizer-BioNTech and Oxford-AstraZeneca vaccines on covid-19 related symptoms, hospital admissions, and mortality in older adults in England: test negative case-control study. BMJ. 2021 May 13;373:1088.

7. Tetteh JNA, Nguyen VK, Hernandez-Vargas EA. Network models to evaluate vaccine strategies towards herd immunity in COVID-19. J Theor Biol. 2021 Dec 21;531:110894.

8. Tatapudi H, Das R, Das TK. Impact of vaccine prioritization strategies on mitigating COVID-19: an agent-based simulation study using an urban region in the United States. BMC Med Res Methodol. 2021 Dec 5;21(1):272.

9. Foy BH, Wahl B, Mehta K, Shet A, Menon GI, Britto C. Comparing COVID-19 vaccine allocation strategies in India: A mathematical modelling study. Int J Infect Dis. 2021 Feb;103:431–8.

10. Borchering RK, Viboud C, Howerton E, Smith CP, Truelove S, Runge MC, et al. Modeling of Future COVID-19 Cases, Hospitalizations, and Deaths, by Vaccination Rates and Nonpharmaceutical Intervention Scenarios - United States, April-September 2021. MMWR Morb Mortal Wkly Rep. 2021 May 14;70(19):719–24.

11. Hammond J, Leister-Tebbe H, Gardner A, Abreu P, Bao W, Wisemandle W, et al. Oral Nirmatrelvir for High-Risk, Nonhospitalized Adults with Covid-19. N Engl J Med. 2022 Apr 14;386(15):1397–408.

12. Shah MM, Joyce B, Plumb ID, Sahakian S, Feldstein LR, Barkley E, et al. Paxlovid Associated with Decreased Hospitalization Rate Among Adults with COVID-19 - United States, April-September 2022. MMWR Morb Mortal Wkly Rep. 2022 Dec 2;71(48):1531–7.

13. Gottlieb RL, Vaca CE, Paredes R, Mera J, Webb BJ, Perez G, et al. Early Remdesivir to Prevent Progression to Severe Covid-19 in Outpatients. N Engl J Med. 2022 Jan 27;386(4):305–15.

14. Dougan M, Azizad M, Chen P, Feldman B, Frieman M, Igbinadolor A, et al. Bebtelovimab, alone or together with bamlanivimab and etesevimab, as a broadly neutralizing monoclonal antibody treatment for mild to moderate, ambulatory COVID-19 [Internet]. medRxiv; 2022 [cited 2023 Feb 8]. p. 2022.03.10.22272100. Available from: https://www.medrxiv.org/content/10.1101/2022.03.10.22272100v1

15. Ison MG, Hayden FG. Antiviral Agents Against Respiratory Viruses. Infectious Diseases. 2017;1318–1326.e2.

16. Wikle NB, Tran TNA, Gentilesco B, Leighow SM, Albert E, Strong ER, et al. SARS-CoV-2 epidemic after social and economic reopening in three U.S. states reveals shifts in age structure and clinical characteristics. Science Advances. 2022 Jan 26;8(4):eabf9868.

17. Tran TNA, Wikle NB, Yang F, Inam H, Leighow S, Gentilesco B, et al. SARS-CoV-2 Attack Rate and Population Immunity in Southern New England, March 2020 to May 2021. JAMA Netw Open. 2022 May 26;5(5):e2214171.

18. Tran TNA, Wikle NB, Albert E, Inam H, Strong E, Brinda K, et al. Optimal SARS-CoV-2 vaccine allocation using real-time attack-rate estimates in Rhode Island and Massachusetts. BMC Med. 2021 Jul 13;19(1):162.

19. Rhode Island COVID-19 Data FAQ [Internet]. [cited 2023 Feb 7]. Available from: https://ridoh-covid-19-data-faq-rihealth-rihealth.hub.arcgis.com/

20. COVID-19 Reported Patient Impact and Hospital Capacity by State Timeseries | HealthData.gov [Internet]. [cited 2023 Feb 7]. Available from: https://healthdata.gov/Hospital/COVID-19-Reported-Patient-Impact-and-Hospital-Capa/g62h-syeh

21. COVID-19 Vaccinations in the United States,Jurisdiction | Data | Centers for Disease Control and Prevention [Internet]. [cited 2023 Feb 7]. Available from: https://data.cdc.gov/Vaccinations/COVID-19-Vaccinations-in-the-United-States-Jurisdi/unsk-b7fc

22. Influenza Vaccination Coverage for All Ages (6+ Months) | Data | Centers for Disease Control and Prevention [Internet]. [cited 2023 Feb 7]. Available from: https://data.cdc.gov/Flu-Vaccinations/Influenza-Vaccination-Coverage-for-All-Ages-6-Mont/vh55-3he6

23. COVID-19 Therapeutics Thresholds, Orders, and Replenishment by Jurisdiction | HHS/ASPR [Internet]. [cited 2023 Feb 7]. Available from: https://aspr.hhs.gov:443/COVID-19/Therapeutics/Orders/Pages/default.aspx

24. Boucau J, Uddin R, Marino C, Regan J, Flynn JP, Choudhary MC, et al. Characterization of virologic rebound following nirmatrelvir-ritonavir treatment for COVID-19. Clin Infect Dis. 2022 Jun 1;ciac512.

25. Charness ME, Gupta K, Stack G, Strymish J, Adams E, Lindy DC, et al. Rebound of SARS-CoV-2 Infection after Nirmatrelvir-Ritonavir Treatment. N Engl J Med. 2022 Sep 15;387(11):1045–7.

26. Coulson JM, Adams A, Gray LA, Evans A. COVID-19 “Rebound” associated with nirmatrelvir/ritonavir pre-hospital therapy. Journal of Infection. 2022 Oct 1;85(4):436–80.

27. Ranganath N, O’Horo JC, Challener DW, Tulledge-Scheitel SM, Pike ML, O’Brien M, et al. Rebound Phenomenon After Nirmatrelvir/Ritonavir Treatment of Coronavirus Disease 2019 (COVID-19) in High-Risk Persons. Clinical Infectious Diseases. 2022 Jun 14;ciac481.

28. Wang Y, Chen X, Xiao W, Zhao D, Feng L. Rapid COVID-19 rebound in a severe COVID-19 patient during 20-day course of Paxlovid. Journal of Infection. 2022 Nov 1;85(5):e134–6.

29. Wang L, Berger NA, Davis PB, Kaelber DC, Volkow ND, Xu R. COVID-19 rebound after Paxlovid and Molnupiravir during January-June 2022 [Internet]. Infectious Diseases (except HIV/AIDS); 2022 Jun [cited 2023 Jan 3]. Available from: http://medrxiv.org/lookup/doi/10.1101/2022.06.21.22276724

30. Li H, Gao M, You H, Zhang P, Pan Y, Li N, et al. Association of Nirmatrelvir/Ritonavir Treatment on Upper Respiratory Severe Acute Respiratory Syndrome Coronavirus 2 Reverse Transcription-Polymerase Chain Reaction (SARS-Cov-2 RT-PCR) Negative Conversion Rates Among High-Risk Patients With Coronavirus Disease 2019 (COVID-19). Clinical Infectious Diseases. 2022 Jul 23;ciac600.

31. Pfizer. Pfizer’s Novel COVID-19 Oral Antiviral Treatment Candidate Reduced Risk of Hospitalization or Death by 89% in Interim Analysis of Phase 2/3 EPIC-HR Study. Nota de prensa. 2021;

32. Andrews N, Stowe J, Kirsebom F, Toffa S, Rickeard T, Gallagher E, et al. Covid-19 Vaccine Effectiveness against the Omicron (B.1.1.529) Variant. N Engl J Med. 2022 Apr 21;386(16):1532–46.

33. Ferdinands JM, Rao S, Dixon BE, Mitchell PK, DeSilva MB, Irving SA, et al. Waning of vaccine effectiveness against moderate and severe covid-19 among adults in the US from the VISION network: test negative, case-control study. BMJ. 2022 Oct 3;379:e072141.

34. Tartof SY, Slezak JM, Fischer H, Hong V, Ackerson BK, Ranasinghe ON, et al. Effectiveness of mRNA BNT162b2 COVID-19 vaccine up to 6 months in a large integrated health system in the USA: a retrospective cohort study. The Lancet. 2021 Oct;398(10309):1407–16.

35. De Giorgi V, West KA, Henning AN, Chen LN, Holbrook MR, Gross R, et al. Naturally Acquired SARS-CoV-2 Immunity Persists for Up to 11 Months Following Infection. The Journal of Infectious Diseases. 2021 Oct 28;224(8):1294–304.

36. Ssentongo P, Ssentongo AE, Voleti N, Groff D, Sun A, Ba DM, et al. SARS-CoV-2 vaccine effectiveness against infection, symptomatic and severe COVID-19: a systematic review and meta-analysis. BMC Infectious Diseases. 2022 May 7;22(1):439.

37. Bobrovitz N, Ware H, Ma X, Li Z, Hosseini R, Cao C, et al. Protective effectiveness of previous SARS-CoV-2 infection and hybrid immunity against the omicron variant and severe disease: a systematic review and meta-regression. Lancet Infect Dis. 2023 Jan 18;S1473-3099(22)00801-5.

38. Hansen CH, Michlmayr D, Gubbels SM, Mølbak K, Ethelberg S. Assessment of protection against reinfection with SARS-CoV-2 among 4 million PCR-tested individuals in Denmark in 2020: a population-level observational study. Lancet. 2021 Mar 27;397(10280):1204–12.

39. Nordström P, Ballin M, Nordström A. Risk of SARS-CoV-2 reinfection and COVID-19 hospitalisation in individuals with natural and hybrid immunity: a retrospective, total population cohort study in Sweden. Lancet Infect Dis. 2022 Jun;22(6):781–90.

40. Lewnard JA, Liu VX, Jackson ML, Schmidt MA, Jewell BL, Flores JP, et al. Incidence, clinical outcomes, and transmission dynamics of severe coronavirus disease 2019 in California and Washington: prospective cohort study. BMJ. 2020 May 22;369:m1923.

41. Wu Y, Kang L, Guo Z, Liu J, Liu M, Liang W. Incubation Period of COVID-19 Caused by Unique SARS-CoV-2 Strains: A Systematic Review and Meta-analysis. JAMA Netw Open. 2022 Aug 22;5(8):e2228008.

42. Boucau J, Marino C, Regan J, Uddin R, Choudhary MC, Flynn JP, et al. Duration of Shedding of Culturable Virus in SARS-CoV-2 Omicron (BA.1) Infection. N Engl J Med. 2022 Jul 21;387(3):275–7.

43. Lewnard JA, Hong VX, Patel MM, Kahn R, Lipsitch M, Tartof SY. Clinical outcomes associated with SARS-CoV-2 Omicron (B.1.1.529) variant and BA.1/BA.1.1 or BA.2 subvariant infection in Southern California. Nat Med. 2022 Sep;28(9):1933–43.

44. COVID-19 Nursing Home Data - Centers for Medicare & Medicaid Services Data [Internet]. [cited 2023 Jan 8]. Available from: https://data.cms.gov/covid-19/covid-19-nursing-home-data

45. Nyberg T, Ferguson NM, Nash SG, Webster HH, Flaxman S, Andrews N, et al. Comparative Analysis of the Risks of Hospitalisation and Death Associated with SARS-CoV-2 Omicron (B.1.1.529) and Delta (B.1.617.2) Variants in England. SSRN Journal [Internet]. 2022 [cited 2023 Jan 3]; Available from: https://www.ssrn.com/abstract=4025932

46. Servadio JL, Thai PQ, Choisy M, Boni MF. Repeatability and timing of tropical influenza epidemics [Internet]. medRxiv; 2022 [cited 2023 Jan 24]. p. 2022.11.04.22281944. Available from: https://www.medrxiv.org/content/10.1101/2022.11.04.22281944v1

47. CDC. Burden of Influenza [Internet]. Centers for Disease Control and Prevention. 2022 [cited 2023 Jan 10]. Available from: https://www.cdc.gov/flu/about/burden/index.html

48. Lopez Bernal J, Andrews N, Gower C, Gallagher E, Simmons R, Thelwall S, et al. Effectiveness of Covid-19 Vaccines against the B.1.617.2 (Delta) Variant. New England Journal of Medicine. 2021 Aug 12;385(7):585–94.

49. Wang L, Volkow ND, Davis PB, Berger NA, Kaelber DC, Xu R. COVID-19 rebound after Paxlovid treatment during Omicron BA.5 vs BA.2.12.1 subvariant predominance period [Internet]. Infectious Diseases (except HIV/AIDS); 2022 Aug [cited 2023 Jan 3]. Available from: http://medrxiv.org/lookup/doi/10.1101/2022.08.04.22278450

50. Jayk Bernal A, Gomes da Silva MM, Musungaie DB, Kovalchuk E, Gonzalez A, Delos Reyes V, et al. Molnupiravir for Oral Treatment of Covid-19 in Nonhospitalized Patients. N Engl J Med. 2022 Feb 10;386(6):509–20.

51. Bubar KM, Reinholt K, Kissler SM, Lipsitch M, Cobey S, Grad YH, et al. Model-informed COVID-19 vaccine prioritization strategies by age and serostatus. Science. 2021 Feb 26;371(6532):916–21.

52. Matrajt L, Eaton J, Leung T, Brown ER. Vaccine optimization for COVID-19: Who to vaccinate first? Sci Adv. 2020 Feb;7(6):eabf1374.

53. Leung K, Jit M, Leung GM, Wu JT. The allocation of COVID-19 vaccines and antivirals against emerging SARS-CoV-2 variants of concern in East Asia and Pacific region: A modelling study. The Lancet Regional Health - Western Pacific. 2022 Apr 1;21:100389.

54. McCreary EK, Bariola JR, Minnier TE, Wadas RJ, Shovel JA, Albin D, et al. The comparative effectiveness of COVID-19 monoclonal antibodies: A learning health system randomized clinical trial. Contemp Clin Trials. 2022 Aug;119:106822.

55. WHO Solidarity Trial Consortium. Remdesivir and three other drugs for hospitalised patients with COVID-19: final results of the WHO Solidarity randomised trial and updated meta-analyses. Lancet. 2022 May 21;399(10339):1941–53.

56. Matrajt L, Brown ER, Cohen MS, Dimitrov D, Janes H. Could widespread use of antiviral treatment curb the COVID-19 pandemic? A modeling study.

57. Respiratory Syncytial Virus (RSV) | NIH: National Institute of Allergy and Infectious Diseases [Internet]. [cited 2023 Jan 23]. Available from: https://www.niaid.nih.gov/diseases-conditions/respiratory-syncytial-virus-rsv

58. Underlying Cause of Death, 1999-2020 Request [Internet]. [cited 2023 Jan 23]. Available from: https://wonder.cdc.gov/ucd-icd10.html

59. Loomba S, de Figueiredo A, Piatek SJ, de Graaf K, Larson HJ. Measuring the impact of COVID-19 vaccine misinformation on vaccination intent in the UK and USA. Nat Hum Behav. 2021 Mar;5(3):337–48.

60. Whitehead HS, French CE, Caldwell DM, Letley L, Mounier-Jack S. A systematic review of communication interventions for countering vaccine misinformation. Vaccine. 2023 Jan 27;41(5):1018–34.

61. Joshi A, Kaur M, Kaur R, Grover A, Nash D, El-Mohandes A. Predictors of COVID-19 Vaccine Acceptance, Intention, and Hesitancy: A Scoping Review. Front Public Health. 2021;9:698111.

62. Andreasen V. Dynamics of annual influenza A epidemics with immuno-selection. J Math Biol. 2003 Jun;46(6):504–36.

63. Andreasen V. The Final Size of an Epidemic and Its Relation to the Basic Reproduction Number. Bull Math Biol. 2011 Oct 1;73(10):2305–21.

64. Johnson BT, Carey MP, Marsh KL, Levin KD, Scott-Sheldon LAJ. Interventions to reduce sexual risk for the human immunodeficiency virus in adolescents, 1985-2000: a research synthesis. Arch Pediatr Adolesc Med. 2003 Apr;157(4):381–8.

65. Liu Y, Morgenstern C, Kelly J, Lowe R, CMMID COVID-19 Working Group, Jit M. The impact of non-pharmaceutical interventions on SARS-CoV-2 transmission across 130 countries and territories. BMC Med. 2021 Feb 5;19(1):40.

66. Boehmer TK. Racial and Ethnic Disparities in Outpatient Treatment of COVID-19 - United States, January-July 2022. MMWR Morb Mortal Wkly Rep [Internet]. 2022 [cited 2023 Jan 24];71. Available from: https://www.cdc.gov/mmwr/volumes/71/wr/mm7143a2.htm

67. Persad G, Peek ME, Shah SK. Fair Allocation of Scarce Therapies for Coronavirus Disease 2019 (COVID-19). Clin Infect Dis. 2022 Aug 24;75(1):e529–33.

68. Archive: COVID-19 Vaccination and Case Trends by Age Group, United States | Data | Centers for Disease Control and Prevention [Internet]. [cited 2023 Feb 5]. Available from: https://data.cdc.gov/Vaccinations/Archive-COVID-19-Vaccination-and-Case-Trends-by-Ag/gxj9-t96f

69. Wong GLH, Yip TCF, Lai MSM, Wong VWS, Hui DSC, Lui GCY. Incidence of Viral Rebound After Treatment With Nirmatrelvir-Ritonavir and Molnupiravir. JAMA Netw Open. 2022 Dec 1;5(12):e2245086.

70. Deo R, Choudhary MC, Moser C, Ritz J, Daar ES, Wohl DA, et al. Viral and Symptom Rebound in Untreated COVID-19 Infection. medRxiv. 2022 Aug 2;2022.08.01.22278278.

71. Soares H, Baniecki M, Cardin RD, Leister-Tebbe H, Zhu Y, Guan S, et al. Viral Load Rebound in Placebo and Nirmatrelvir-Ritonavir Treated COVID-19 Patients is not Associated with Recurrence of Severe Disease or Mutations [Internet]. 2022 [cited 2023 Feb 8]. Available from: https://www.researchsquare.com

72. Jochmans D, Liu C, Donckers K, Stoycheva A, Boland S, Stevens SK, et al. The substitutions L50F, E166A and L167F in SARS-CoV-2 3CLpro are selected by a protease inhibitor in vitro and confer resistance to nirmatrelvir [Internet]. bioRxiv; 2022 [cited 2023 Jan 12]. p. 2022.06.07.495116. Available from: https://www.biorxiv.org/content/10.1101/2022.06.07.495116v2

73. Zhou Y, Gammeltoft KA, Ryberg LA, Pham LV, Tjørnelund HD, Binderup A, et al. Nirmatrelvir-resistant SARS-CoV-2 variants with high fitness in an infectious cell culture system. Science Advances. 2022 Dec 21;8(51):eadd7197.

