## Supplemental Materials for "Benefits of near-universal vaccination and treatment access to manage COVID-19 burden in the United States"

S1 Text. Estimating current and future vaccine coverages

To inform our estimates of annual vaccine coverage, we used data on COVID-19 vaccinations from CDC (21) between December 1, 2021 and November 30, 2022 (Table S3, Fig S3). In these data, the timing of individual-level booster shots is not provided, making it difficult to know exactly how many individuals in the US received at least one dose of a COVID-19 vaccine during this time period. To account for this, we generated a range of vaccine coverage estimates, based on conservative and optimistic assumptions about which fields represent duplicate doses (i.e., multiple doses were given to the same individual during our one-year period).

1. Conservative: this estimate assumes that all individuals competing their two-course series received the first dose of this series during the time period, and that all second boosters were given to individuals who received a first booster during the time period (i.e., $d_{cons} = d_{admin} - d_{series} - d_{second}$). In a population of nearly 332 million, this yields a conservative coverage estimate of 36%.

2. Optimistic: this estimate assumes that the only duplicate doses were those individuals completing their two-course series (i.e., $d_{opt} = d_{admin} - d_{series}$). This yields an optimistic coverage estimate of 49%.

Future trends of COVID-19 vaccination uptake are assumed to follow influenza vaccination. The average proportion of vaccines administered in each month was calculated, then linearly interpolated to generate weekly estimates of vaccination rates. Because the age groups (6 months – 4 years-old; 5-12 years-old, 13-17 years-old, 18-49 years-old, 50-64 years-old, and 65 years-old and above) in the data are different from the model, the age-specific number of new vaccinees were calculated from the coverage rate and the US census in 2020, and were allocated to the age groups in our model under an assumption of uniformity within each of the above age brackets. The weekly relative coverage rate was calculated by dividing the coverage rate this week by the total coverage achieved in the year. The age-specific annual relative coverage rate is used as the future trend of COVID-19 vaccination coverage.

Table S1: Updated clinical parameters for the Omicron variant. Clinical parameters retained from prior time periods have been previous published (17).

| Parameter | Pre-Omicron | Omicron |
|---|---|---|
| Incubation period | 6 days (fixed) (17) | 3.42 days (41) |
| Infectious period | 5 days (fixed) (17) | 6 days (42) |
| Probability of hospitalization by age | 0 (0-9 years),<br>0.012 (10-19 years),<br>0.021 (20-29 years),<br>0.030 (30-39 years),<br>0.048 (40-49 years),<br>0.078 (50-59 years),<br>0.147 (60-69 years),<br>0.285 (70-79 years),<br>0.314 (>=80 years) (fitted) (17) | 0 (0-9 years),<br>0.005 (10-19 years),<br>0.021 (20-29 years),<br>0.024(30-39 years),<br>0.021 (40-49 years),<br>0.038 (50-59 years),<br>0.056 (60-69 years),<br>0.180 (70-79 years),<br>0.242 (>= 80 years) (43) |
| Mean length of stay in hospital | 10.7 days (fixed) (17) | 13.3 days (43) |
| Probability of ICU admission in hospital | 0.304 (0-9 years),<br>0.293 (10-19 years),<br>0.283 (20-29 years),<br>0.301 (30-39 years),<br>0.463 (40-49 years),<br>0.4245 (50-59 years),<br>0.46 (60-69 years),<br>0.484 (70-79 years),<br>0.416 (>= 80 years) (fixed) (40) | 0.152 (0-9 years),<br>0.1465 (10-19 years),<br>0.141 (20-29 years),<br>0.151 (30-39 years),<br>0.232 (40-49 years),<br>0.212 (50-59 years),<br>0.23 (60-69 years),<br>0.242 (70-79 years),<br>0.208 (>= 80 years) (40) |
| Probability of ventilation in ICU | 0.66 (fitted) (16) | 0.238 (43) |
| Probability of death in non-ICU hospital care | 0 (0-69 years) (fixed),<br>0.025 (70-79 years),<br>0.050 (>= 80 years) (fitted) (17) | 0 (0-69 years),<br>0.005 (70-79 years),<br>0.011 (>= 80 years) (43) |
| Probability of death outside of hospital | 0 (0-59) (fixed),<br>0.013 (60-69 years),<br>0.042 (70-79 years),<br>0.227 (>= 80 years) (fitted) (17) | 0 (0-59 years),<br>0.0013 (60-69 years),<br>0.0061(70-79 years),<br>0.074 (>= 80 years) (44,45) |

Table S2. Treatment efficacies of COVID-19 therapeutics

| Treatment | Efficacy | Reference |
|---|---|---|
| Nirmatrelvir/ritonavir (Paxlovid) | 87.8 reduction of severity compared to unvaccinated population (prospective study) | (11) |
| | 50% reduction in hospitalization compared to a population mixed with unvaccinated (15%) and vaccinated (85%) individuals (retrospective study) | (12) |
| Molnupiravir (Lagevrio) | 29.9% reduction of hospitalization or death compared to unvaccinated population (prospective study) | (50) |
| Remdesivir | 86.8% reduction of hospitalization or death compared to unvaccinated population (prospective study) | (13) |
| Bebtelovimab | 36-40% reduction of hospitalization compared to a population mixed with unvaccinated and vaccinated individuals (prospective study) | (14) |

Table S3: COVID-19 vaccine doses administered by age group in the United States between December 1, 2021-November 30, 2022.

| | Value |
|---|---|
| Total doses administered, $d_{admin}$ | 193M |
| Total first doses administered, $d_{first}$ | 34M |
| Total completed two-course series, $d_{series}$ | 31M |
| Total "additional" doses administered (non-bivalent), $d_{additional}$ | 73M |
| Total second boosters administered (non-bivalent), $d_{second}$ | 41M |
| Total bivalent boosters administered, $d_{bivalent}$ | 40M |

Table S4. Estimated durations of immunity following vaccination or infection from literature.

| ref | Article | Duration | % immune (inf) | % immune (vax) | Location | Time | Notes |
|---|---|---|---|---|---|---|---|
| (32) | Andrews et al, 2022 (NEJM) | 2-4 weeks | | 82.8 | England | Nov 2020 - Jan 2022 | ChAdOx1 against Delta |
| | | 25+ weeks | | 43.5 | | | |
| | | 20-24 weeks | | none | | | ChAdOx1 against Omicron |
| | | 2-4 weeks | | 65.5 | | | |
| | | 15-19 weeks | | 15.4 | | | |
| | | 25+ weeks | | 8.8 | | | mRNA-1273 against omicron |
| | | 2-4 weeks | | 75.1 | | | |
| | | 25+ weeks | | 14.9 | | | |
| | | 2-4 weeks | | 62.4 | | | BNT162b2 booster after ChAdOx1 |
| | | 10+ weeks | | 39.6 | | | |
| | | 2-4 weeks | | 70.1 | | | mRNA-1273 booster after ChAdOx1 |
| | | 5-9 weeks | | 60.9 | | | |
| | | 5-9 weeks | | 46.7 | | | ChAdOx1 booster after ChAdOx1 |
| | | 2-4 weeks | | 67.2 | | | BNT162b2 booster after BNT162b2 |
| | | 10+ weeks | | 45.7 | | | |
| | | 2-4 weeks | | 73.9 | | | mRNA booster after BNT162b2 |
| | | 5-9 weeks | | 64.4 | | | |
| | | 2-4 weeks | | 64.9 | | | BNT162b2 booster after mRNA-1273 |
| | | 2-4 weeks | | 66.3 | | | mRNA-1273 booster after mRNA-1273 |
| (33) | | 4-6 months | | 86 | | | |

| | | | | | | | |
|---|---|---|---|---|---|---|---|
| | Ferdinands et al., 2022 (BMJ) | 6-8 months | | 79 | United States | Jan 2021 - Jul 2022 | VE of 2 doses against ED or urgent care visit against Delta |
| | | 10-12 months | | 66 | | | |
| | | 4-6 month | | 88 | | | VE of 3 doses against ED or urgent care visit against Delta |
| | | 4-6 month | | 37 | | | VE of 2 doses against ED or urgent care visit against Omicron |
| | | 6-8 months | | 30 | | | |
| | | 10-12 months | | 35 | | | |
| | | 12-14 months | | 16 | | | |
| | | 16-18 months | | 22 | | | |
| | | 4-6 months | | 46 | | | VE of 3 doses against ED or urgent care visit against Omicron |
| | | 6-8 months | | 26 | | | |
| | | 8+ months | | 17 | | | |
| (34) | Tartof et al., 2021 (Lancet) | 5+ months | | 47 | United States | Dec 2020 - Aug 2021 | VE of fully vaccinated against infection, 12+ |
| | | | | 43 | | | VE of fully vaccinated against infection, 65+ |
| | | 4+ months | | 53 | | | VE of fully vaccinated against Delta |
| | | | | 67 | | | VE of fully vaccinated against non-Delta |
| (35) | De Giorgi et al., 2021 (JID) | 11 months | 63 | | United States | Apr 2020 - Feb 2021 | Detectable neutralizing titers |
| (36) | | 4 months | | 71.4 | International | | |

| | | | | | | | |
|---|---|---|---|---|---|---|---|
| | Ssentongo et al., 2022 (BMCID) | 5 months | | 21.8 | | Dec 2019 - Nov 2021 (published dates) | Systematic Review and meta analysis (18 articles) |
| (37) | Bobrovitz et al., 2023 (Lancet ID) | 3 months | 65.2 | | International | Jan 2020- Jun 2022 (publication date) | Systematic review and meta analysis (11 studies) |
| | | 12 months | 24.7 | | | | |
| | | 15 months | 15.5 | | | | |
| | | 3 months | 69 | | | | Hybrid immunity with primary series vax |
| | | 12 months | 41.8 | | | | Hybrid immunity with primary series vax |
| | | 3 months | 68.6 | | | | Hybrid immunity with booster |
| | | 6 months | 46.5 | | | | Hybrid immunity with booster |
| (38) | Hansen et al. 2021 (Lancet) | 3-6 months | 80.5 | | Denmark | Sep - Dec 2020 | Compared rates of Covid among those with and without infection before June 2020 |
| (39) | Nordstrom et al., 2022 (Lancet ID) | 3-6 months | 0.9696 | | Sweden | March 2020 - Oct 2021 | Infection vs no immunity |
| | | 6-9 months | 92 | | | | |
| | | 2+ months | | 45 | | | one dose hybrid vs natural infection only |
| | | 2+ months | | 56 | | | two dose hybrid vs natural infection only |

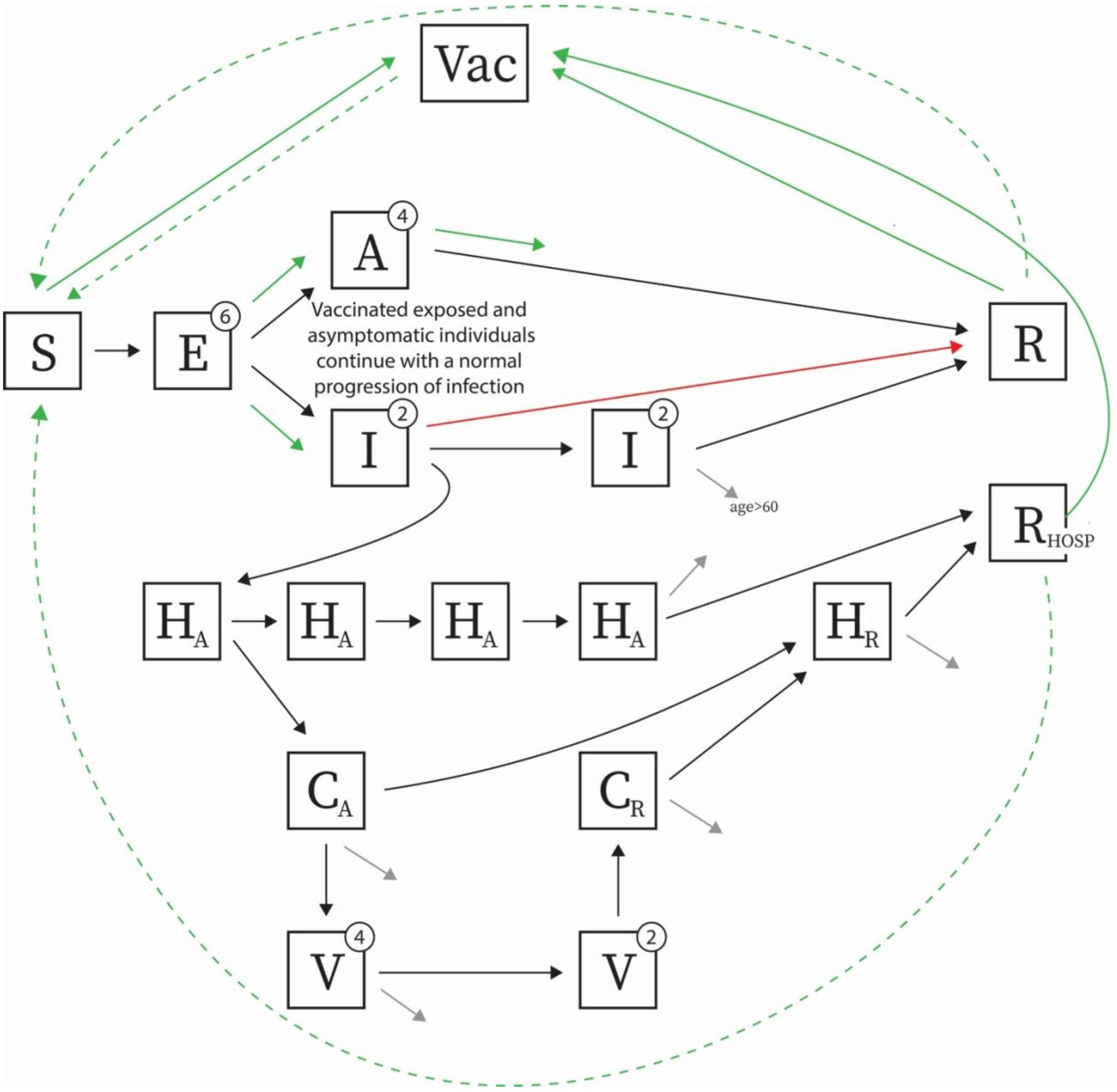


Figure S1. Model diagram. Adapted from previously published model (17). Changes made include adding a one-stage vaccine compartment (Vac), waning of infection-induced and vaccine-induced immunity, and fast recovery from infection. The green solid line indicates the vaccine-seeking behavior, while only susceptible and recovered individuals will benefit from vaccination. The green dashed line indicates the waning immunity acquired from infection or vaccination. The red solid line indicates the fast recovery of the infected individuals after successful treatment. The infected individuals with the failed treatment continue a normal progress of infection but with lower risk of hospitalization.

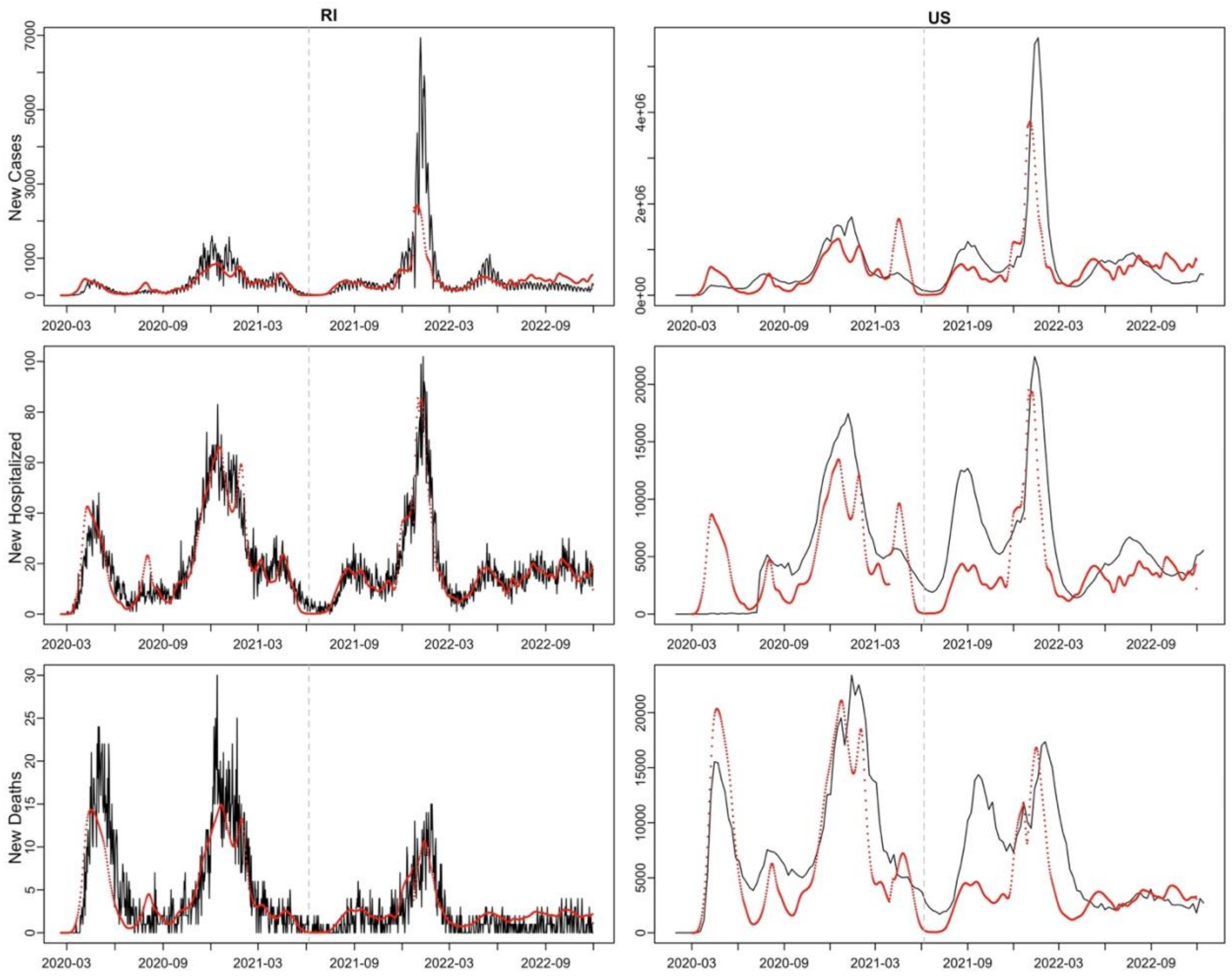


Figure S2. Calibrated output (red points) compared to the observed data (black line) in RI (left) and the US (right). After June 6, 2021 (grey vertical dashed line), the population mixing rates were calibrated to the hospitalized data in RI. New cases and new deaths in RI were simulated based on calibrated population mixing rates. The new cases, hospitalizations, and deaths in the US were scaled up from RI using the ratio of the total cases in different variant-dominant period between RI and the US. The scaled-up outputs for the US match the trends in the observed data.

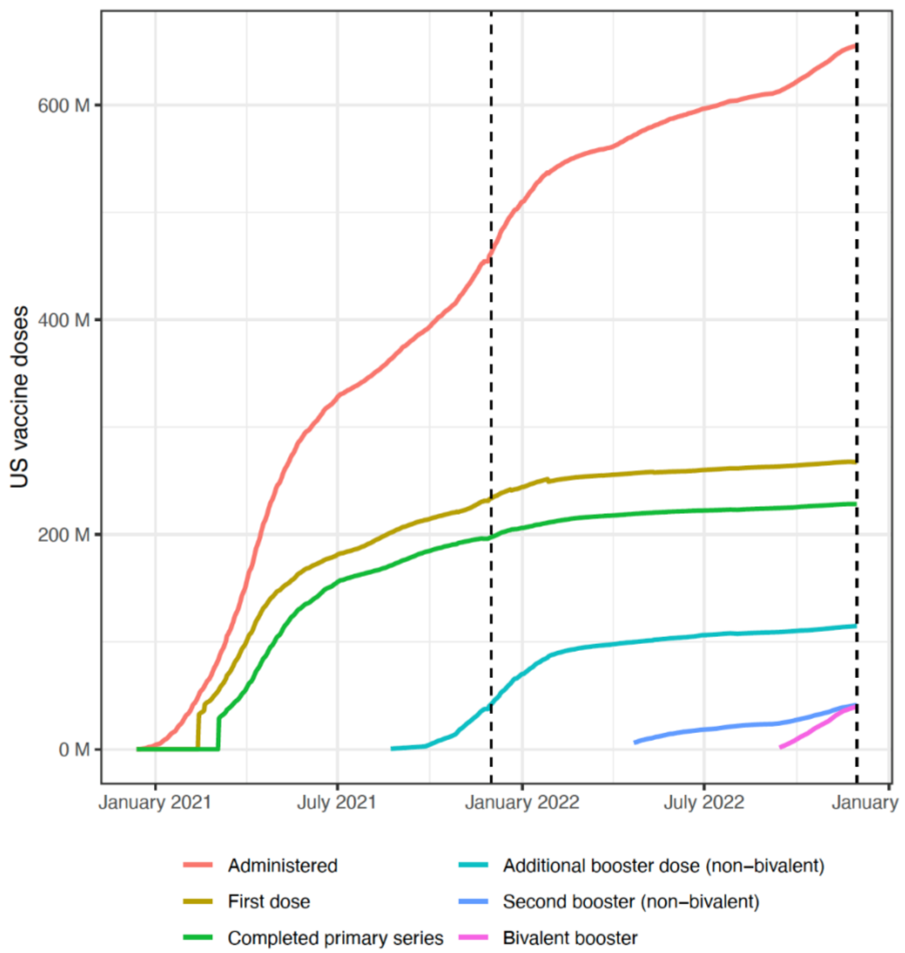


Figure S3. Doses of COVID-19 vaccines administered in the United States, including total doses administered, number of the first and second doses of the primary two-dose series administered, non-bivalent booster doses, second non-bivalent booster doses, and bivalent booster doses. Coverage between December 1, 2021 and November 30, 2022 (vertical dashed lines) were applied to model parameterizations.

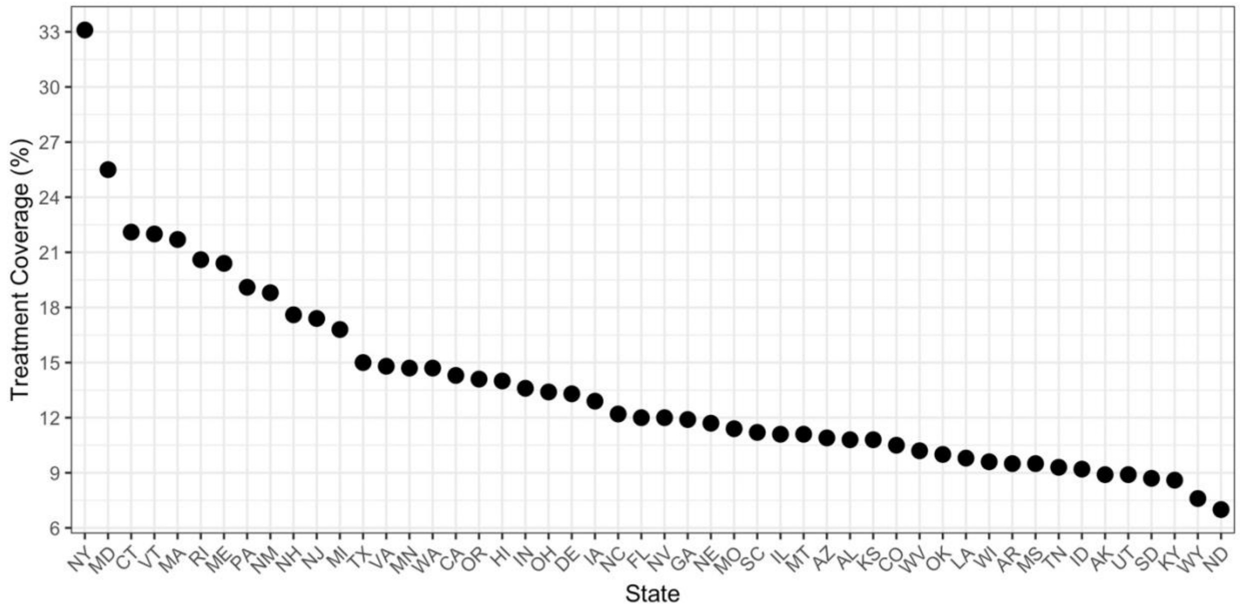


Figure S4. Coverage of Paxlovid in 50 states as of Dec 11, 2022. This is calculated as the cumulative administered courses of Paxlovid on Dec 11, 2022, divided by the number of patients from Jan 1, 2022, to Dec 11, 2022.

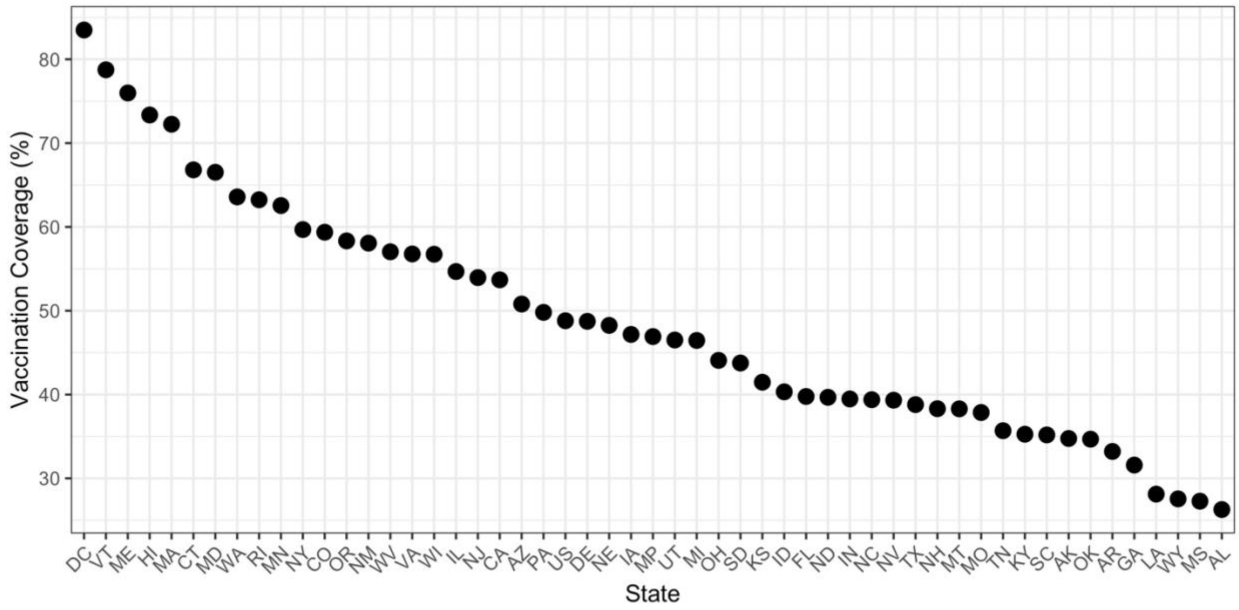


Figure S5. Vaccination coverage in 50 states between Dec 1, 2021, and Nov 30, 2022. Coverage is calculated as the number of administered doses of either a two-course primary series or booster divided by state-wide population (age > 6 months).

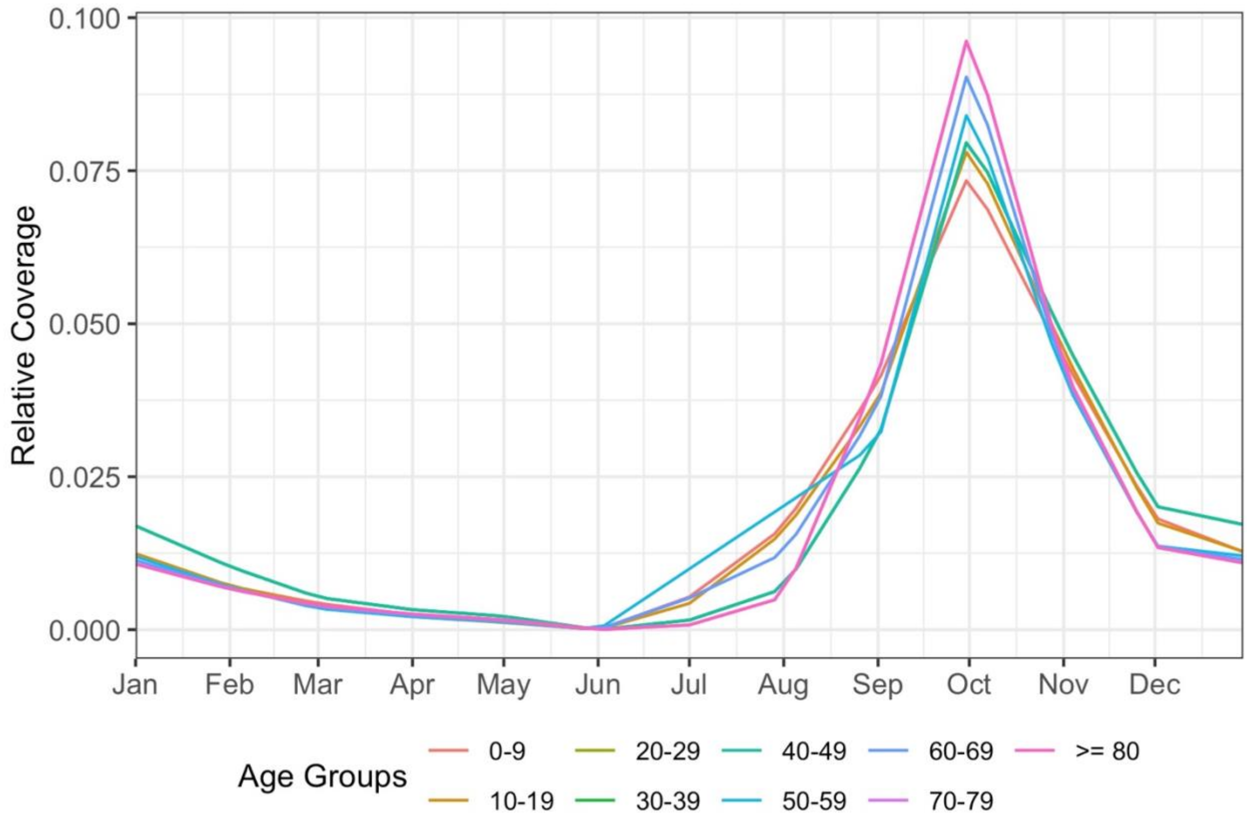


Figure S6. Weekly percentages of achieved coverage of influenza vaccination by age groups by calendar month. The trends observed were applied to COVID-19 vaccination administration in model projections.

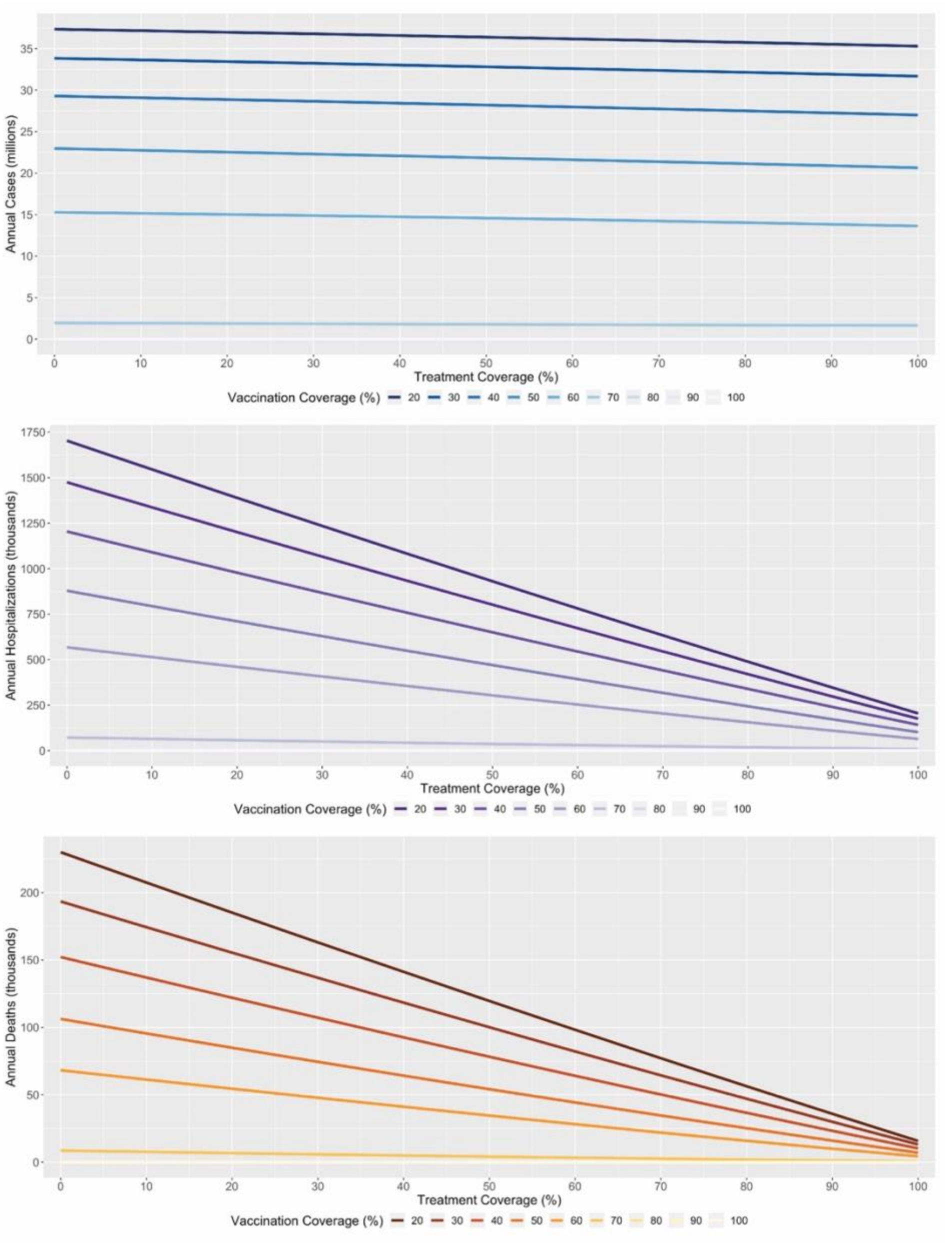


Figure S7. Burden reduction slopes following treatment coverage under different vaccination coverages. Reductions in hospitalizations and deaths are more pronounced under low-vaccination circumstances.

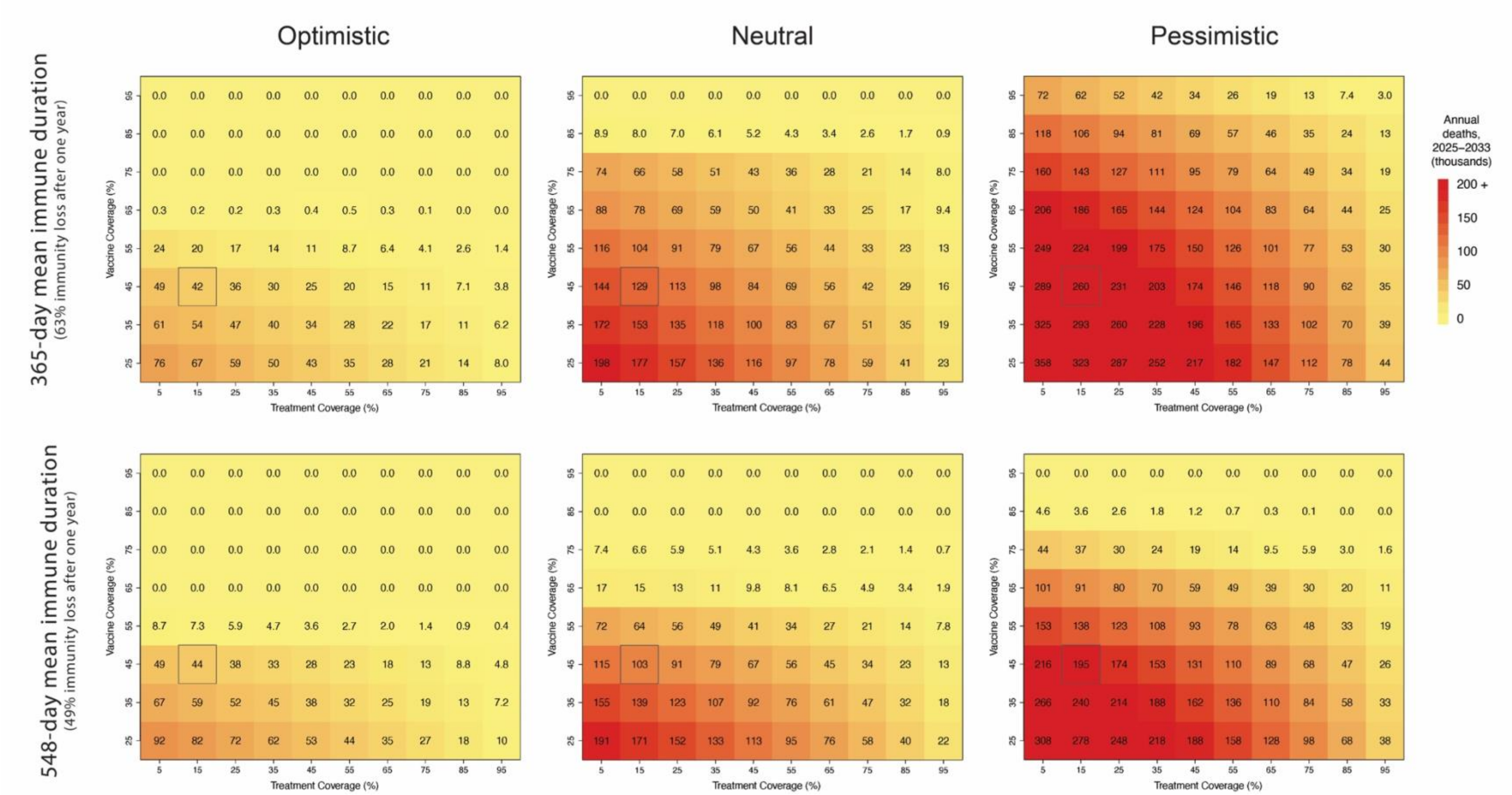


Figure S8. Heatmaps of annual mortality under combinations of vaccine and treatment coverage under each transmission scenario and rate of immune waning.

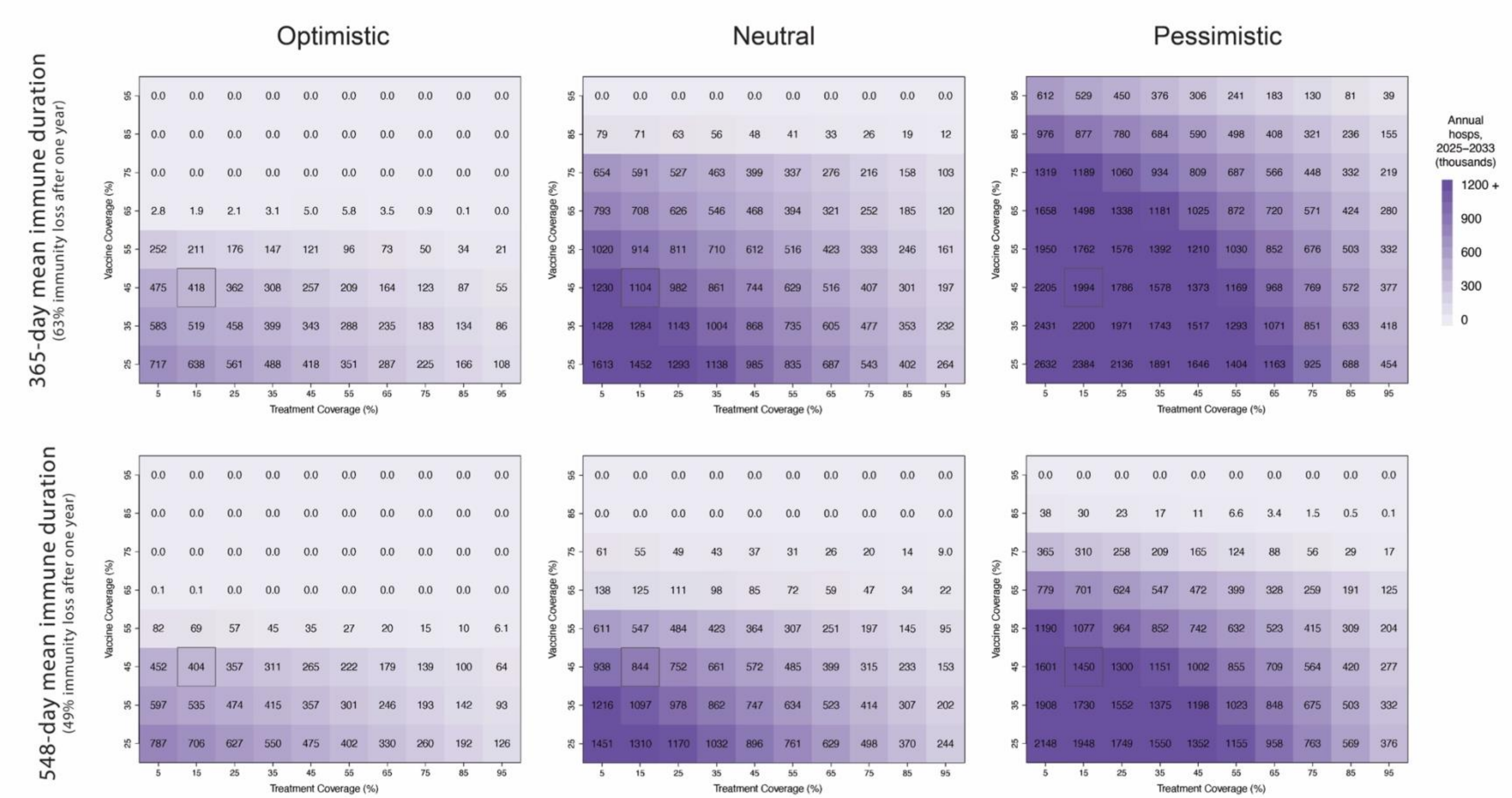


Figure S9. Heatmaps of annual hospitalizations under combinations of vaccine and treatment coverage under each transmission scenario and rate of immune waning.

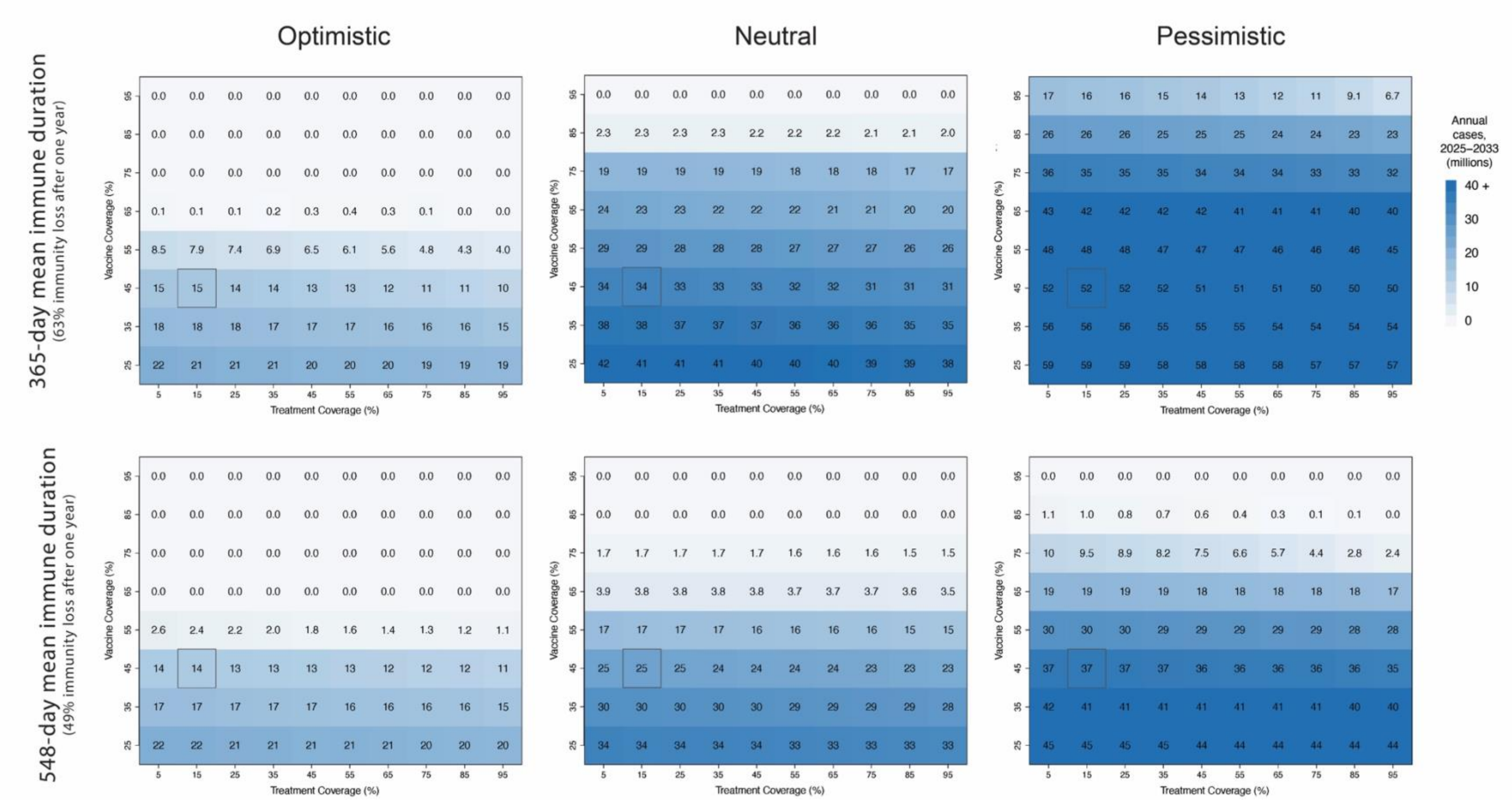


Figure S10. Heatmaps of annual incident cases under combinations of vaccine and treatment coverage under each transmission scenario and rate of immune waning.

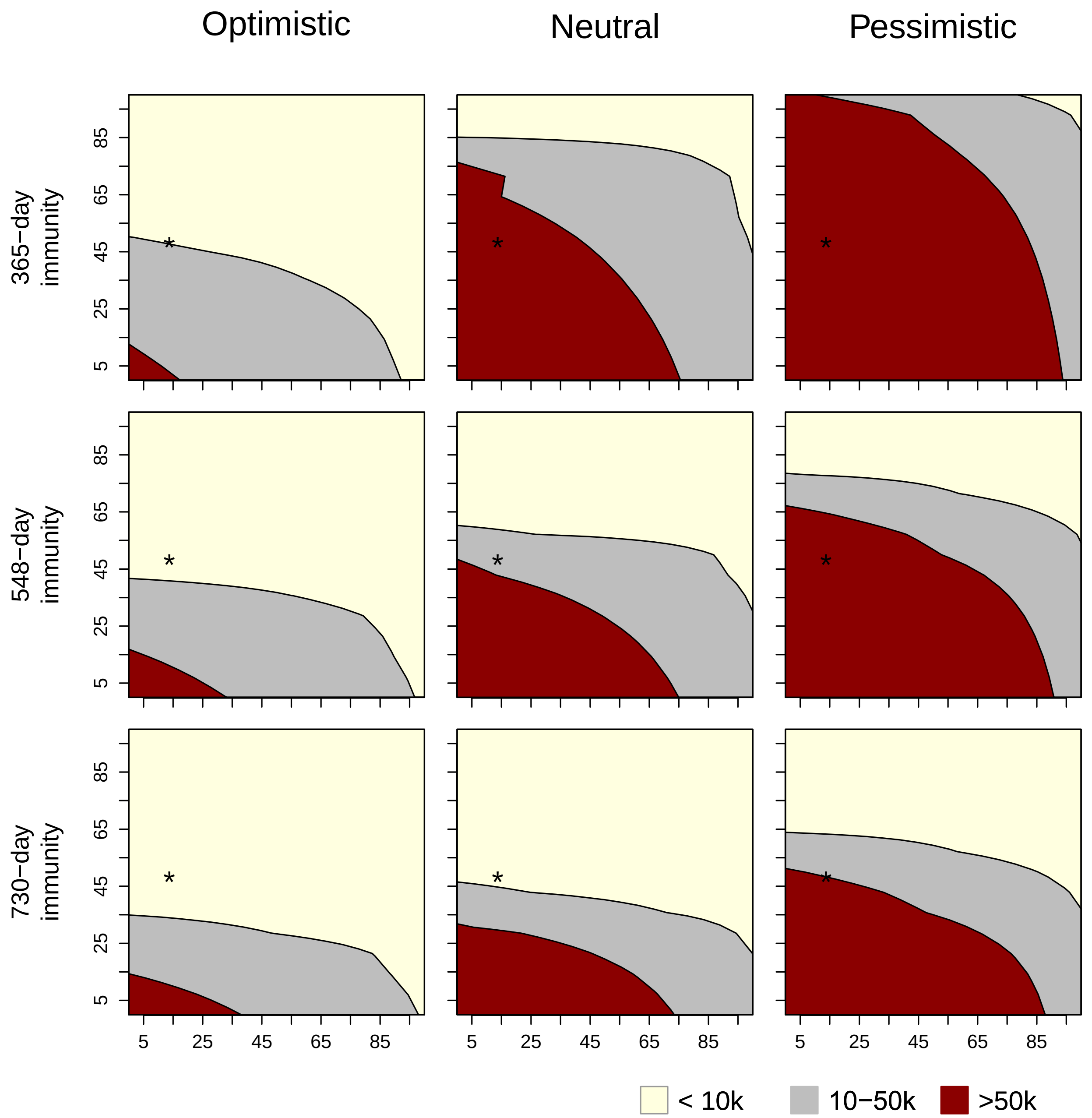


Figure S11. Combinations of treatment coverage (horizontal axis) and vaccine coverage (vertical axis) that lead to COVID-19 mortality within the range of annual influenza mortality (10,000 – 50,000 deaths) as well as below or over this range. The starred point in the plots represents the current treatment and vaccine coverage in the United States.

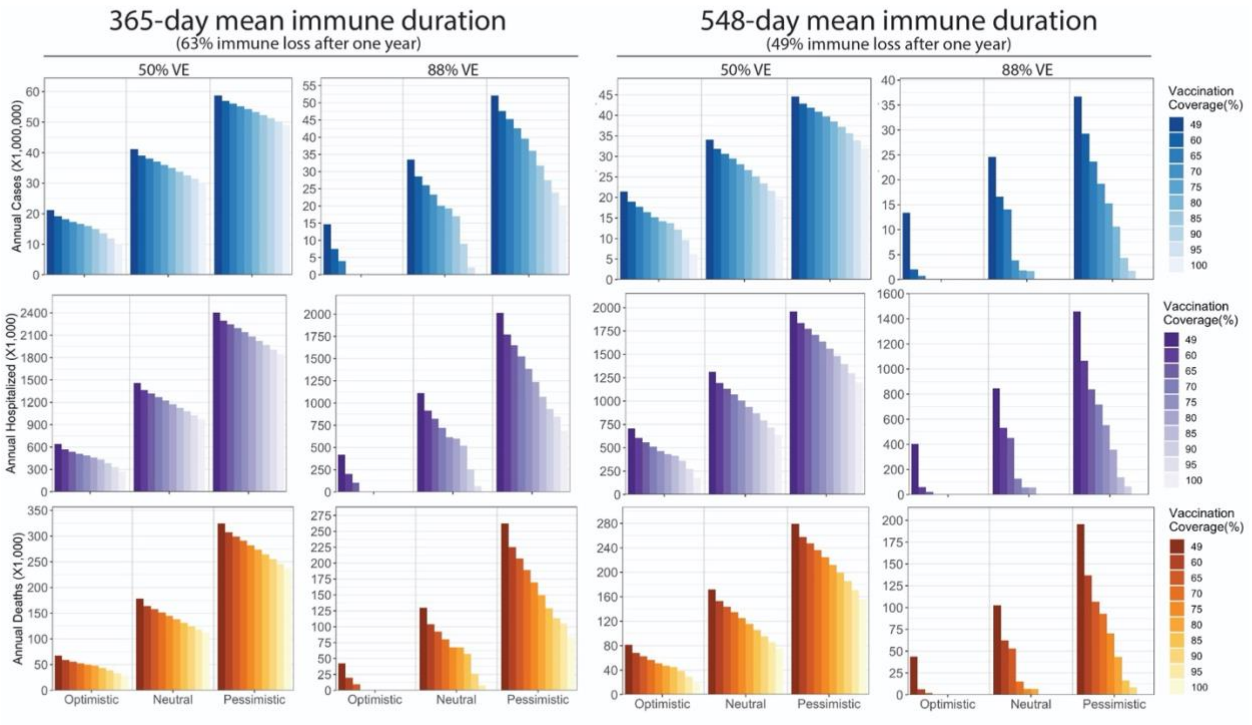


Figure S12. Annual burden reduction between 2025 and 2033 given 50% and 88% vaccine effectiveness. The treatment coverage in the entire period is 13.7%.

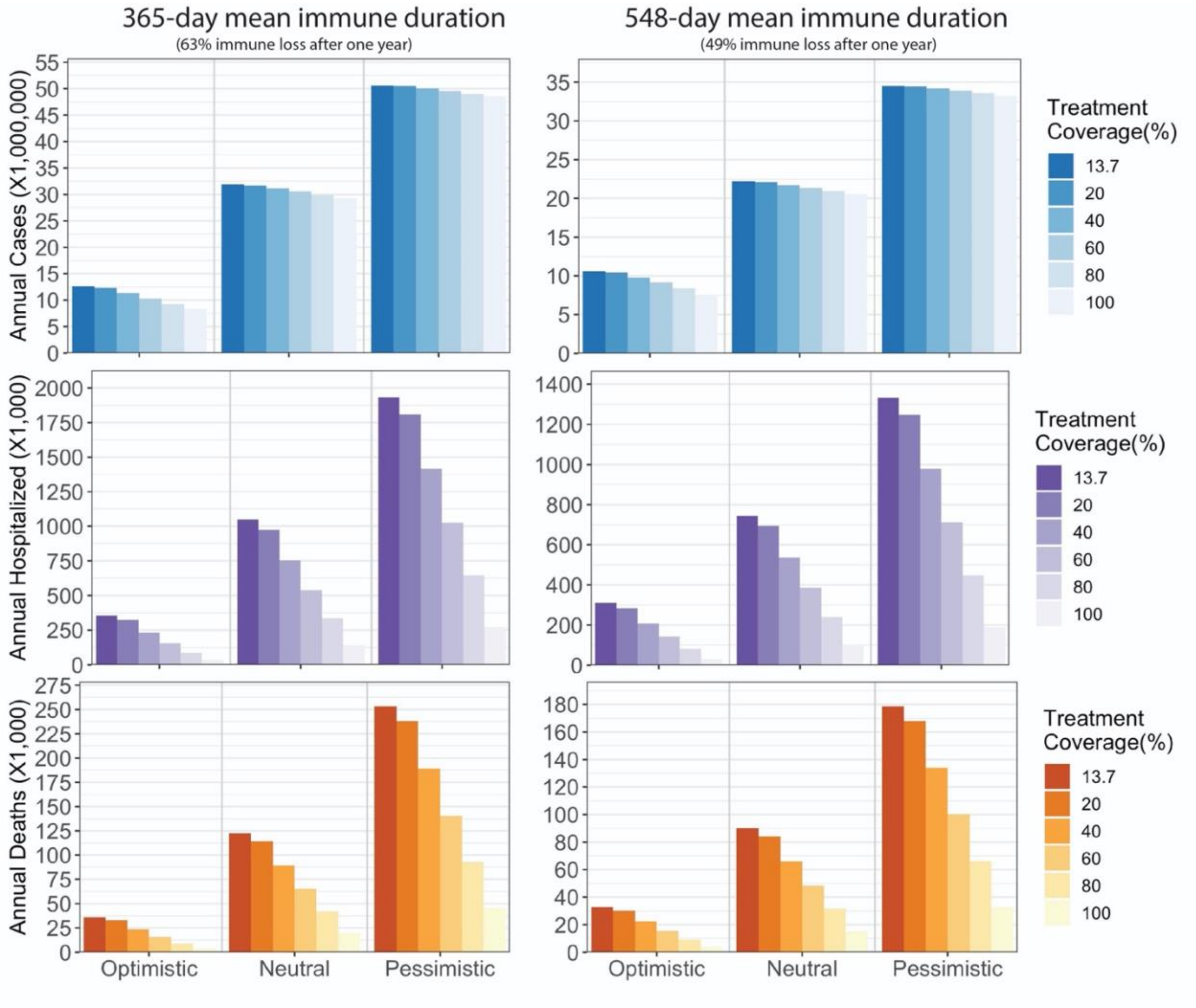


Figure S13. Annual burden reduction between 2025 and 2033 given 20% probability of treatment failure. Vaccination coverage during the entire period is 49%.

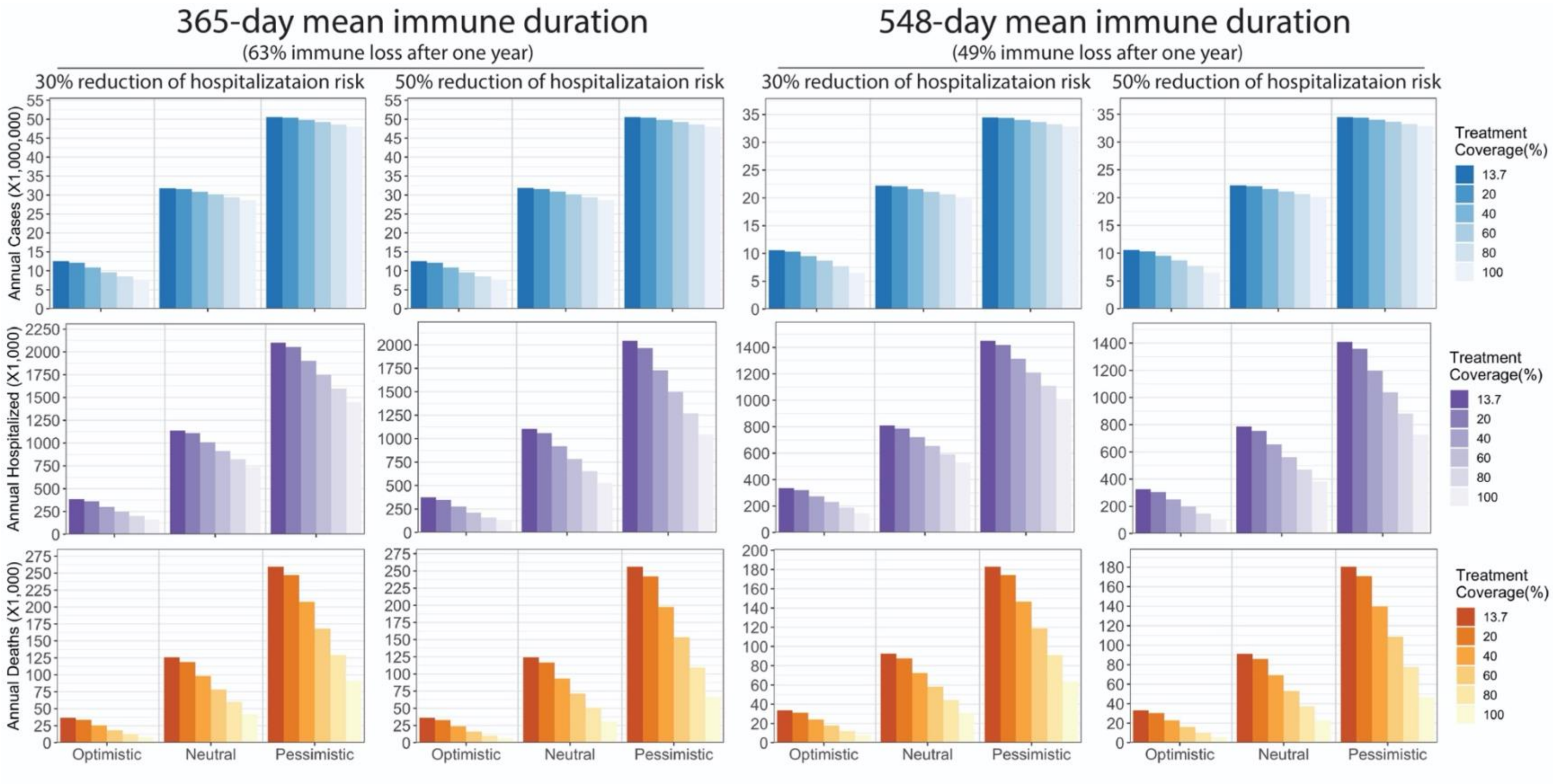


Figure S14. Annual burden reduction between 2025 and 2033 given 30% and 50% risk reduction to hospitalization after failed treatment. The probability of failed treatment is 0.035. Vaccination coverage during the entire period is 49%.
